# Triage-driven diagnosis for early detection of esophageal cancer using deep learning

**DOI:** 10.1101/2020.07.16.20154732

**Authors:** Marcel Gehrung, Mireia Crispin-Ortuzar, Adam G. Berman, Maria O’Donovan, Rebecca C. Fitzgerald, Florian Markowetz

**Affiliations:** Cancer Research UK Cambridge Institute, University of Cambridge, UK; The Alan Turing Institute, London, UK; MRC Cancer Unit, University of Cambridge, UK; Department of Pathology, Cambridge University Hospitals NHS Trust, UK

## Abstract

Deep learning methods have been shown to achieve excellent performance on diagnostic tasks, but it is still an open challenge how to optimally combine them with expert knowledge and existing clinical decision pathways. This question is particularly important for the early detection of cancer, where high volume workflows might potentially benefit substantially from automated analysis. Here, we present a deep learning framework to analyse samples of the Cytosponge®-TFF3 test, a minimally invasive alternative to endoscopy, for detecting Barrett’s Esophagus, the main precursor of esophageal cancer. We trained and independently validated the framework on data from two clinical trials, analysing a combined total of 4,662 pathology slides from 2,331 patients. Our approach exploits screening patterns of expert gastrointestinal pathologists and established decision pathways to define eight triage classes of varying priority for manual expert review. By substituting manual review with automated review in low-priority classes, we can reduce pathologist workload by up to 66% while matching the diagnostic performance of expert pathologists. These results lay the foundation for tailored, semi-automated decision support systems embedded in clinical workflows.

## Introduction

Early detection of cancer often leads to better survival (*1*), because pre-malignant lesions and early stage tumors can be more effectively treated (*2*). Most pre-malignant lesions amenable to early detection rely on targeted sampling and show only minor tissue changes on pathology assessment (*3–5*). In addition, pathology procedures often involve laborious and time-consuming steps which can lead to errors and adversely affect patient care (*6*). Recent developments in Artificial Intelligence (AI) have achieved excellent performance on diagnostic tasks (*7–9*). However, understanding how these techniques can be integrated into clinical workflows most efficiently and to assess the actual benefits they bring remains a challenge. The design of a clinical decision support system needs to balance its performance against workload reduction and potential economic impact. Replacing pathologists entirely could lead to substantial workload reduction, but such an approach would only be viable if performance remains comparable to that of human experts. Between a fully automated approach and the *status quo* of fully manual review lies a semi-automated approach, which uses computational methods to triage patients and only presents pathologists with difficult cases. A semi-automated approach will not reduce workload as much as a fully automated approach, but its performance benefits from existing expert knowledge and heuristics. Here we present such a semi-automated triage system using deep learning for the early detection of esophageal cancer.

Esophageal cancer is the sixth most common cause for cancer related deaths (*10*). Patients usually present at an advanced stage with dysphagia and weight loss, and the 5-year overall survival of esophageal adenocarcinoma (EAC), one of two pathological subtypes, is 13% (*11*). EAC can arise from a precursor lesion called Barrett’s Esophagus (BE) (*12, 13*), providing an effective starting point for early detection. BE occurs in patients with Gastresophageal Reflux Disease (GERD), a digestive disorder where acid and bile from the stomach return into the esophagus leading to heartburn symptoms. In Western countries, 10 to 15% of the adult population are affected by GERD (*14*) and, therefore, at an increased risk of having BE. The pathognomonic feature of BE is intestinal metaplasia (IM), a process whereby the stratified squamous epithelial lining localized in the lower esophagus is replaced with columnar epithelium containing goblet cells (*15, 16*). The conventional diagnosis of BE requires an invasive endoscopic procedure of the upper gastrointestinal tract. However, there is no routine endoscopic screening of the GERD population and thus the vast majority of BE patients are undiagnosed (*14*).

Cytosponge-TFF3 is a non-endoscopic, minimally invasive diagnostic test for BE (*17–19*). It is a cell collection device consisting of a compressed sponge on a string inside a gelatin capsule. The capsule is swallowed by the patient and the gelatin dissolves in the stomach after a few minutes, allowing the sponge to expand. The sponge is then withdrawn from the stomach by the attached string, sampling superficial epithelial cells from the top of the stomach, the esophagus, and the oropharynx (Figure 1a). Therefore, the cellular composition of the sample is dominated by squamous cells, gastric columnar epithelium, and respiratory epithelium as well as any Barrett’s cells, if present. Following removal, the device is placed in a container with preservative solution and the sampled cells are processed, embedded in paraffin and stained with Hematoxylin & Eosin (H&E) as well as immunohistochemically stained with Trefoil Factor 3 (TFF3) (*20*). H&E stains allow the identification and quantification of cellular phenotypes, which is critical for quality control. TFF3 is over-expressed in mucin-producing goblet cells which are a key feature of BE. TFF3 also functions as a protector of the mucosa from insults, stabilizes the mucus layer, and promotes healing of the epithelium (*21*). TFF3 stains allow the identification and quantification of goblet cells, which are indicative of IM. Therefore, TFF3 is the key diagnostic biomarker for BE (*20*).

**Figure 1:**
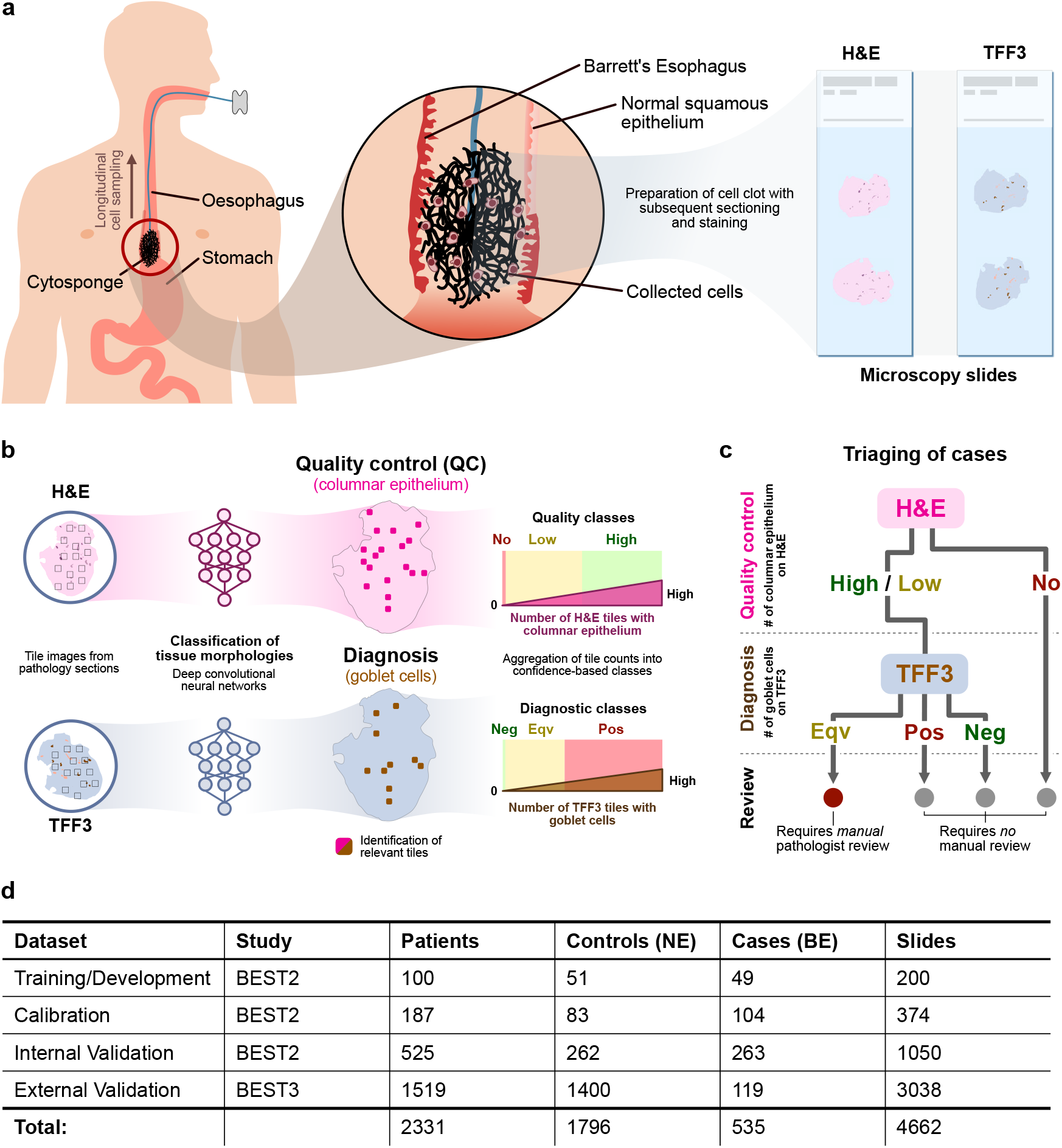
Cytosponge procedure with conceptual patient triage scheme and data summary. **a** During withdrawal the sponge samples superficial epithelial cells from the top of the stomach and the esophagus. These cells are processed into a cell block, then sectioned and stained with Hematoxylin & Eosin (H&E) and Trefoil Factor 3 (TFF3). **b** Convolutional neural networks, trained on an independent training dataset, are used for inference of H&E and TFF3 stains. The resulting tile maps are analysed for relevant regions (columnar epithelium on H&E and goblet cells on TFF3 stain) and aggregated into quality control and diagnostic classes based on tile detections. **c** Quality and diagnostic classes are mapped to a conceptualised pathway for sample stratification. The review layer (bottom) describes to what extent a human pathologist has to review the microscopy slides. (Pos = Positive, Neg = Negative) **d** Overview of data used in this study.

The Cytosponge-TFF3 approach has profound and well-tested clinical significance. It offers, with substantial clinical trial data underpinning its efficacy, a long-awaited diagnostic alternative to endoscopy (BEST1 (*17*), BEST2 (*18*), BEST3 (*22*)). The BEST3 study found that the Cytosponge-TFF3 test had in excess of a 10-fold increase in detection of Barrett’s compared to usual clinical care in which patients with heartburn receive medication and an endoscopy if deemed necessary. This performance makes the Cytosponge a major advance in patient management. The BEST3 study also concluded that the pathology assessment is a major bottleneck for scaling the test to large patient populations. Since the analysis of Cytosponge-TFF3 pathology slides is a very laborious process due to the large amount of sampled cellular material. It comprises several time-consuming tasks such as assessing the amount of sampled material and checking the presence of gastric-type columnar epithelium to confirm that the capsule reached the stomach, followed by assessment for the presence of goblet cells indicative of BE. Though effective, the laboriousness of this process gives rise to a major opportunity for a clinical decision support system to improve analysis and scalability of the Cytosponge-TFF3 test.

Here, we use a deep learning approach for quality control and diagnosis of pathology slides for the Cytosponge-TFF3 test (Figure 1b). We propose a triage-driven approach, which retains diagnostic accuracy by leveraging the decision-making rules of expert gastrointestinal pathologists (Figure 1c). We train, calibrate, and internally validate our approach on data of the BEST2 multi-centre clinical trial (*18*) and externally validate it in an independent cohort from the recent BEST3 multi-centre trial (*22*) (Figure 1d). Additionally, we explore in a simulation study how well our results generalise to more general populations.

## Results

### Deep learning models achieve high performance for tile-level classifications

The first step of our approach is based on the tile-level detection of different classes of cells relevant for quality control and diagnosis of BE. For model development and internal validation, we used 812 Cytosponge-TFF3 patient samples with paired pathology and endoscopy data from the BEST2 clinical case-control study (*18*). Samples were randomly divided into training/development (n=100), calibration (n=187) and internal validation (n=525) sets (Figure 1d). An additional independent dataset (n=1,519) from the BEST3 study was used for external validation of the developed approach.

Training sets of larger size did not improve tile-level accuracy (Figure S1). Training, calibration, and validation sets were kept separate. Endoscopic as well as Cytosponge pathology diagnoses were only unblinded after tile-wise tissue classification models were calibrated and validated, respectively. All training slides were tessellated prior to training: For H&E we derived 193,734 tiles from 100 slides and for TFF3 we derived 235,932 tiles from 100 slides (based on the size of annotated areas, see Methods). All tiles were 200-by-200 µm and all labels were taken from expert slide annotations.

For both quality control (H&E) and diagnostic (TFF3) tasks, we trained several state-of-the-art networks (AlexNet (*23*), DenseNet (*24*), Inception v3 (*25*), ResNet-18 (*26*), SqueezeNet (*27*), and VGG-16 (*28*)) and evaluated their performance on the development datatset. Using individual tiles, we compared tile-level precision and recall for classifying columnar epithelium using the presence of gastric-type cells (on H&E) and positive goblet cells (on TFF3) (Table S1, description in Methods): For gastric-type columnar epithelium, VGG-16, DenseNet and Inception v3 achieved the highest recalls (0.950, 0.947, 0.940, respectively) with consistent precisions (0.843, 0.865, 0.857). For goblet cells, VGG-16, Inception v3, and ResNet-18 achieved the highest recalls (0.919, 0.919, 0.912) with consistent precisions (0.856, 0.856, 0.827). Examples for whole slide images classified positive and negative for quality control and diagnosis are shown in Figure 2a. We also observed a relationship between the tile-level results and the complexity of the applied architectures (Table S1).

**Figure 2:**
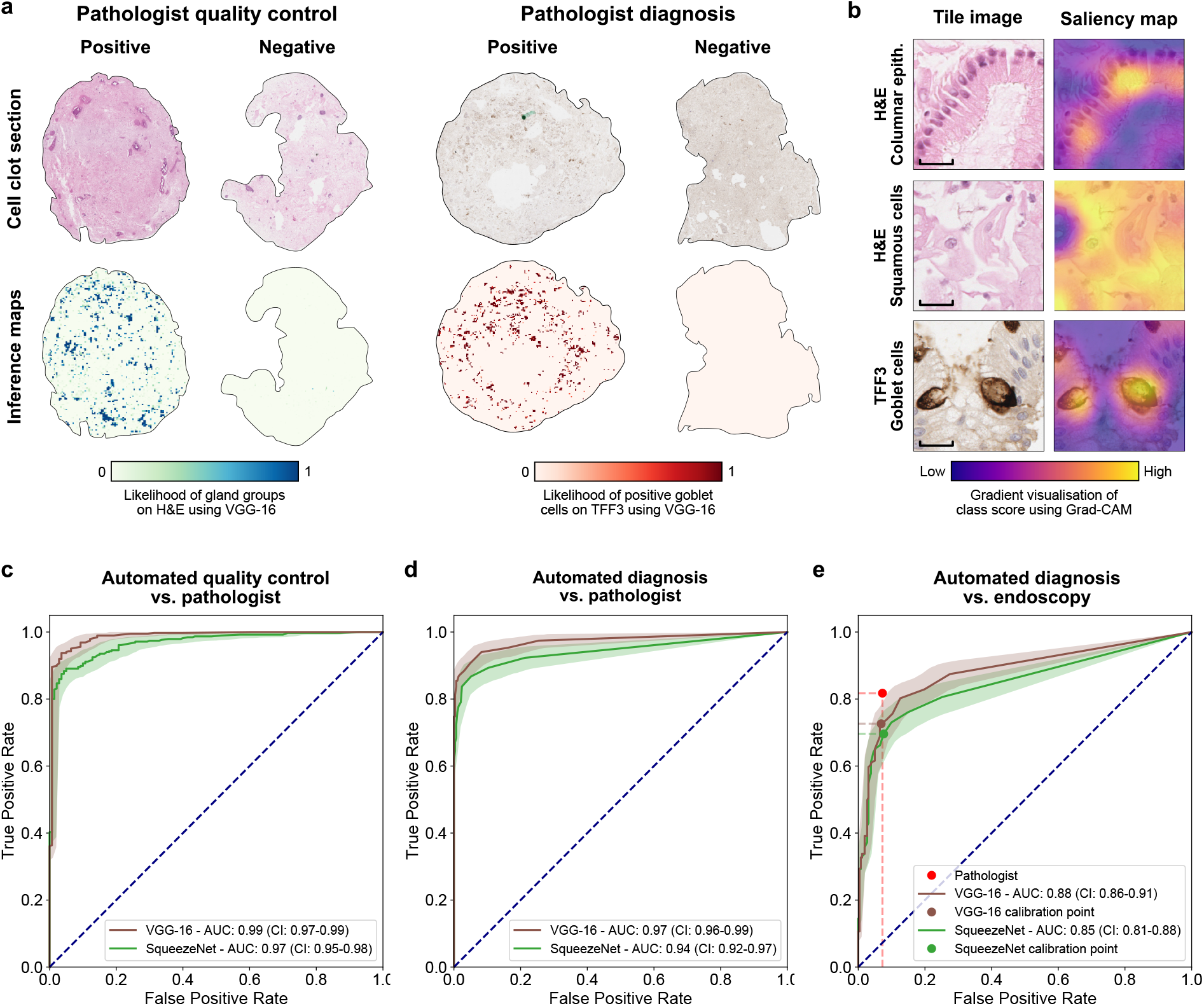
Tile and patient-level classification of Cytosponge-TFF3 samples. **a** Examples of tile-level inference maps for samples, which were classified by a pathologist as positive and negative for quality control (H&E) and diagnosis (TFF3), respectively. **b** Comparison of two tile images from H&E and one tile image from TFF3 with their respective Grad-CAM saliency maps. *Top:* Columnar epithelium (H&E) of gastric type with clear focus on columnar arrangement in saliency map. *Middle:* Squamous cells (H&E) with distributed focus in saliency map. *Bottom:* TFF3-positive goblet cells with localisation in saliency maps. Scale bar = 100 µm. **c** ROC-AUC internal validation cohort analysis of automated tile counts of columnar epithelium on H&E with pathologist ground truth. **d** ROC-AUC internal validation cohort analysis of automated tile counts of positive goblet cells on TFF3 with pathologist ground truth. **e** ROC-AUC internal validation cohort analysis of pathologist and automated tile counts of positive goblet cells on TFF3 with endoscopy ground truth (BE patients defined according to the Prague criteria (Methods) with confirmed IM on biopsy). Pathologist performance: Sensitivity: 81.749% (CI: 76.597% - 85.951%), Specificity: 92.748% (CI: 89.444% - 95.769%). VGG-16 performance (calibration point determined as in Methods): Sensitivity: 72.624% (CI: 67.424% - 78.213%), Specificity: 93.130% (CI: 90.038% - 96.133%). SqueezeNet performance (calibration point, Methods): Sensitivity: 69.582% (CI: 63.915% - 75.522%), Specificity: 92.366% (88.468% - 95.518%). CI = 95% bootstrap confidence interval. Shaded areas show CIs. cohort (Figure 2c-e).

### Saliency maps agree with pathologist criteria for classification of tissue tiles

To understand which characteristics of the tile images were relevant to our models’ classifications, we generated saliency maps using Gradient-weighted Class Activation Mapping (Grad-CAM) (*29*). These maps highlight the local regions of an image most relevant to a model’s identification of a particular class. We generated saliency maps for classes in one H&E-based model (VGG-16) and one TFF3-based model (VGG-16) (Figure 2b). For the gastric-type columnar epithelium class of the H&E-based model, the saliency maps highlight gastric cells by both the linear organisation of their nuclei as well as the presence of a straight border between the cells and the lumen. For the positive class of the TFF3-based model, we found that the saliency maps highlighted the mucin-containing goblet cells that characterise IM with high precision. In addition to the three representative examples in Figure 2b, we compared landmarks selected by an expert pathologist with tile images and respective saliency maps (Figure S2). The saliency maps confirm that the models learned features are similar to those used by pathologists to identify different tissue classes.

### Fully automated approach shows suboptimal performance

Tile-level classifications were aggregated into patient-level classifications using tile counts above thresholds determined by the specificity of expert pathologists on the calibration cohort (Methods, Table S2, Figure S4). We then performed Receiver Operating Characteristics (ROC) analysis with matched Cytosponge pathology and endoscopy ground truth on the internal validation

First, the patient-level scores were compared against the binary Cytosponge-TFF3 ground truth by the pathologist on the internal validation set. For quality control, VGG-16 ranked highest for detecting columnar epithelium in H&E stains (ROC-AUC: 0.99 (CI 95%: 0.98 - 0.99)). SqueezeNet, the least complex architecture we trained, ranked lowest (ROC-AUC: 0.97 (CI 95%: 0.95 - 0.98), Figure 2c). For diagnosis, VGG-16 ranked highest for detecting goblet cells in TFF3 stains (ROC-AUC: 0.97 (CI 95%: 0.96 - 0.99), Figure 2d). Again, SqueezeNet ranked lowest (ROC-AUC: 0.94 (CI 95%: 0.92 - 0.96)). Confidence intervals were derived by bootstrapping (Methods). Results for all architectures are presented in table S3, and fig. S5a/b. In summary, for both quality control and diagnosis in comparison to Cytosponge-TFF3 pathology ground truth, VGG-16 provided the highest performance, and SqueezeNet the lowest.

Next, patient-level counts were compared to endoscopy ground truth for detecting BE on the internal validation set (Methods). This ground truth was defined according to the Prague criteria (Methods) with confirmed IM on endoscopy biopsies (*30*). To calculate sensitivity and specificity for the fully automated method on the internal validation cohort, we used operating points determined on the calibration cohort (Table S2). VGG-16 ranked highest for detecting patients with BE from TFF3 stains (ROC-AUC: 0.88 (CI 95%: 0.85 - 0.91), Sensitivity: 72.62% (CI: 67.42% - 78.21%), Specificity: (93.13% (CI: 90.04% - 96.13%)), Figure 2e). SqueezeNet ranked lowest for detecting patients with BE from TFF3 stains (ROC-AUC: 0.85 (CI 95%: 0.81 - 0.88), Sensitivity: 69.58% (CI: 63.92% - 75.52%), Specificity: 92.37% (88.47% - 95.52%), Figure 2e). For comparison, the pathologists achieve a sensitivity of 81.7% (CI 95%: 77.4% - 86.5%) and a specificity of 92.7% (CI 95%: 89.6% - 95.6%). Performances of all architectures are presented in table S3, and fig. S5c. In summary, results for the fully automated approach on the internal validation cohort showed a loss of sensitivity of 9.1% for BE detection when compared to an expert pathologist.

### Triage-driven approach selects patients for manual review

We then explored whether a different modelling approach based on established decision pathways could boost performance. We developed a triage-driven, semi-automated approach as an alternative to the fully automated approach described above. Both approaches use the same patient-level aggregations as input, but their outputs are different: the fully automated approach tries to directly mimic pathology assessment by classifying patients as positive or negative for BE. In contrast, the triage approach defines different quality and diagnostic confidence classes to select challenging patient samples for manual review. Although it cannot reduce workload as much as a fully automated approach, a triage approach keeps sample stratification more interpretable and transparent.

We first selected deep learning architectures and defined cut-offs for different quality and diagnostic confidence classes based on thresholds determined by two expert observers on the calibration cohort (Figure S6, Methods). For quality confidence classes, pathologists conclude that the sponge reached the stomach if they observe columnar epithelial groups (*18, 20*). We encoded these subjective metrics in a quantitative scheme where the number of tiles detected with gastric-type columnar epithelium on H&E were classified as no confidence, low confidence, or high confidence (Figure S6a, Table S4). For diagnostic confidence classes, the number of tiles detected with TFF3-positive goblet cells were classified as high confidence negative, low confidence equivocal, or high confidence positive (Figure S6b, Table S4). On the internal validation cohort, we observed a visual agreement between these confidence classes and pathology and endoscopy ground truths (Figure 3, Table S5).

**Figure 3:**
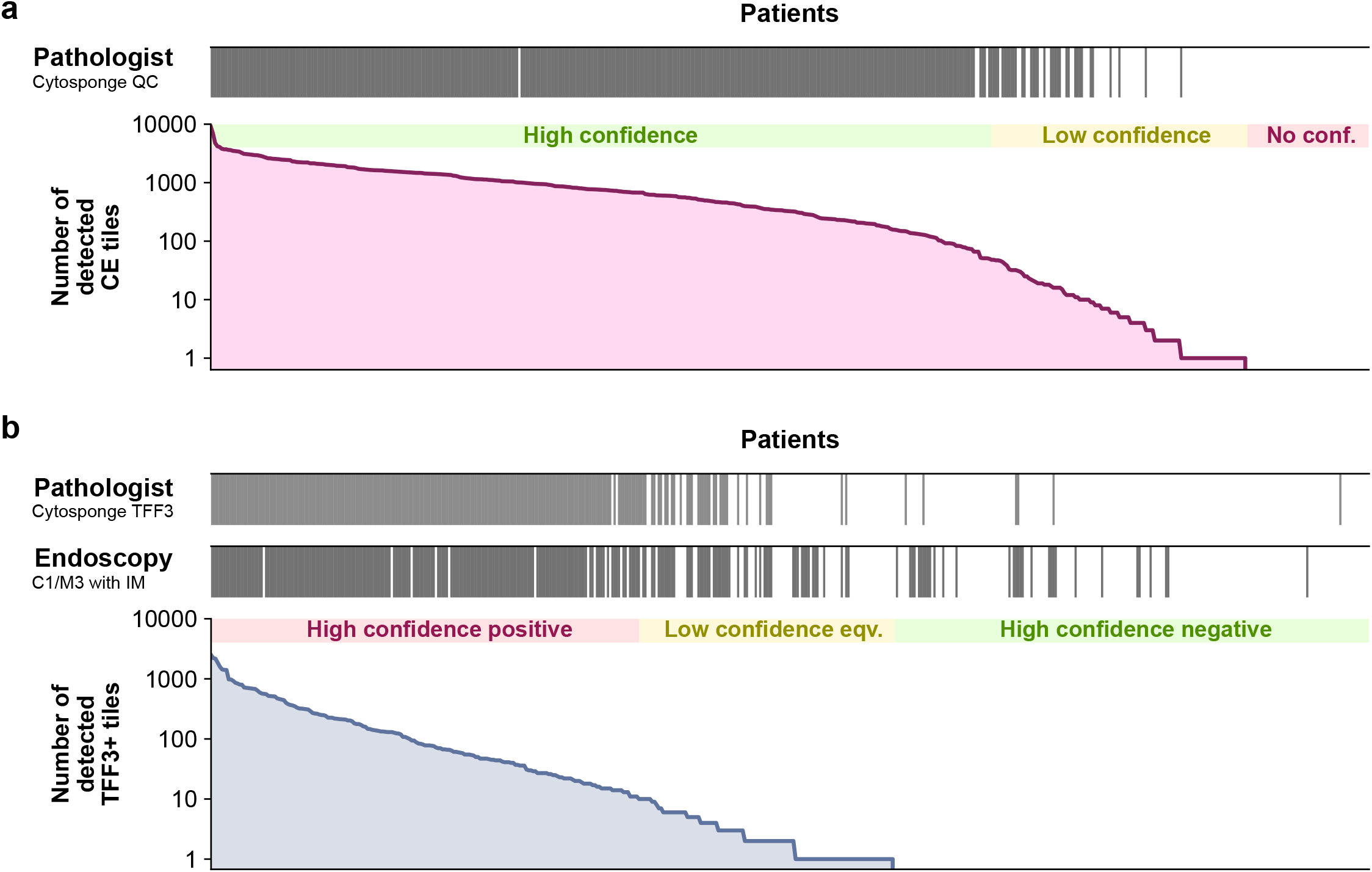
Application of quality control and diagnostic confidence class scheme to internal validation cohort. **a** Quality ground truth by pathologist from Cytosponge (top) compared with number of detected columnar epithelium (CE) tiles on H&E detected by VGG-16 (bottom). **b** Diagnosis ground truth by pathologist from Cytosponge (top), Endoscopy (with confirmed IM on biopsy) ground truth (middle) compared with number of detected TFF3-positive tiles on TFF3 detected by ResNet-18 (bottom) / eqv. = equivocal.

We then combined the quality and diagnostic classes into eight triage classes of varying priority for manual review (Figure 4a). The relative priority of each class was determined by expert pathologists: Cases with low confidence in sample quality (none or few columnar epithelium detected on H&E) or low confidence in diagnosis (few goblet cells detected on TFF3) should be prioritised for human expert assessment over cases with high-confidence positive or negative evidence. In our internal validation cohort, we find that only 13.0% of patients fall into the triage classes with high priority (4 and 5), while 87.0% fall into the other six classes (Figure 4a).

**Figure 4:**
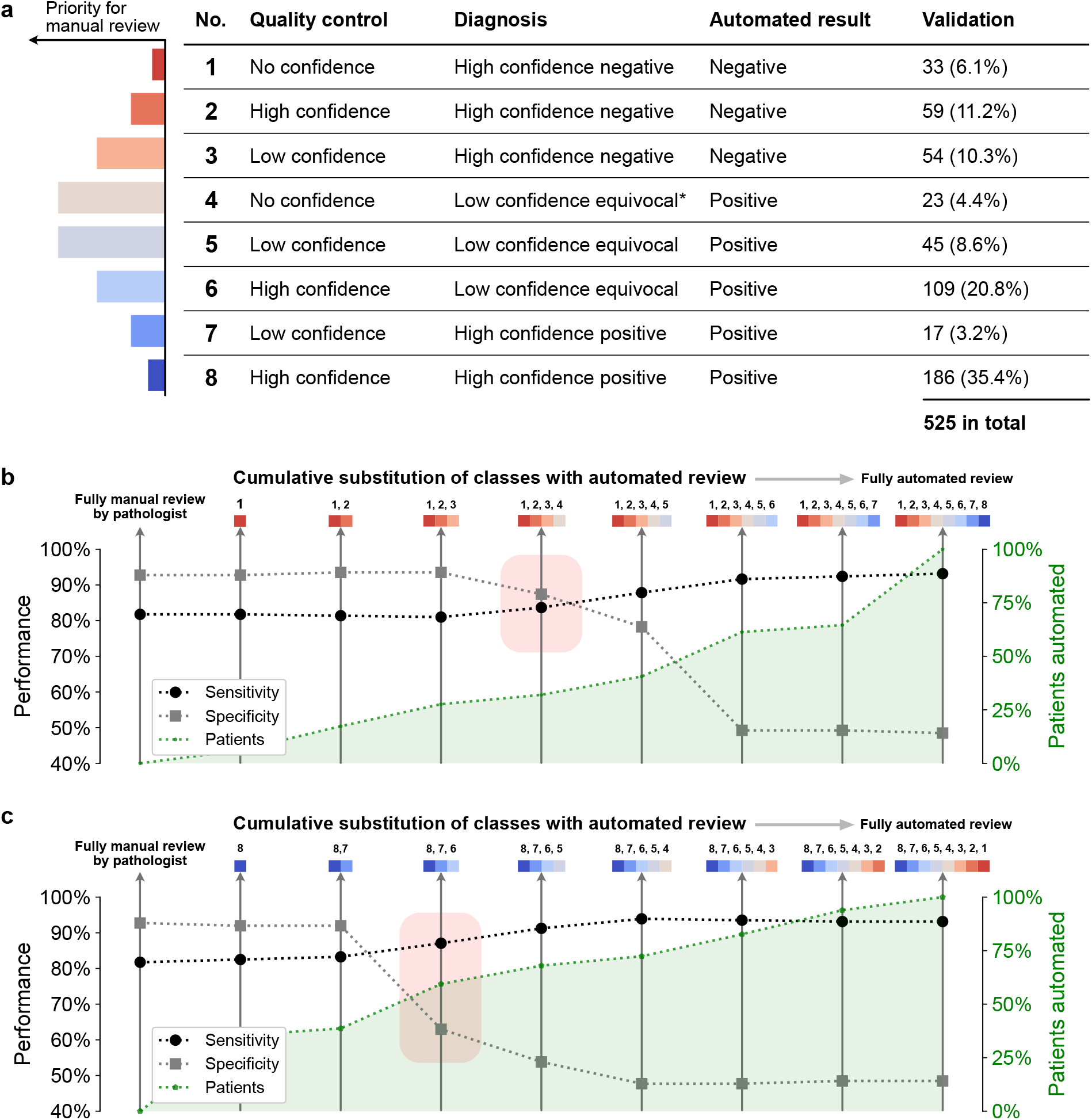
Triage-driven approach with incremental triage class substitution scheme on internal validation set. **a** Table of quality control and diagnosis classes. Each class has been assigned a qualitative priority for manual review. Column ‘Automated result’ refers to the label a sample would be assigned if all samples of this class were automatically reviewed. Asterisk (*): includes combination of no confidence (quality control) and high confidence positive (diagnosis) despite minimal likelihood of occurrence. **b** Cumulative substitution scheme starting with fully manual review, followed by substitution with automated review of class no. 1, then 1 and 2, etc. Red rectangle indicates a drop of performance at substitution stage. **c** Cumulative substitution scheme starting with fully manual review, followed by substitution with automated review of class no. 8, then 8 and 7, etc. Red rectangle indicates a drop of performance at substitution stage.

We next asked which classes can be substituted by automated review while retaining the accuracy of full manual review by a human pathologist (sensitivity: 81.7%; specificity: 92.7%). We applied a cumulative substitution scheme and started by substituting class 1 with automated review, then classes 1 and 2, then classes 1, 2, and 3, and so on. In the validation cohort, we found that sensitivity and specificity remain stable if classes 1, 2, and 3 are substituted, but decrease with the substitution of class 4, 5, and 6 (Figure 4b). Repeating this procedure starting with class 8 shows that sensitivity and specificity are stable if classes 8 or 7 are substituted, but decrease with the substitution of classes 6, 5, and 4 (Figure 4c). These results show that five of the eight classes (1, 2, 3, 7, 8) can be substituted by automated review while three classes (4, 5, 6) should be manually reviewed by a pathologist. This substitution scheme would result in similar performance (sensitivity: 82.5% (CI 95%: 77.3% - 87.2%); specificity: 92.7% (CI 95%: 89.6% - 95.9%)) as fully manual review by a pathologist. These classes cover the majority of patients (66.3% (CI 95%: 62.7% - 70.1%) in validation cohort) and triage-driven, semi automated review would thus save 66% of the pathologists’ workload (Methods) by enabling them to focus on difficult cases while leaving easy cases for automated review.

### Simulation of varying cohort composition corroborates reduction in expected workload

Our case-control cohort is not representative of a real-world population eligible for Cytosponge-TFF3 testing. In our internal validation set we had a disease prevalence of 50.0%, while the prevalence expected in a real-world population with GERD symptoms ranges from 3.0% to 7.5% (*17, 31–33*). Additionally, the allocation of samples to triage classes depends directly on the amount of sampled cellular material and the resulting sample confidence, which can vary widely and might improve with future refinements of the device administration procedure.

To understand how our results generalize, we devised a simulation approach to vary how many samples have BE and how many samples are allocated to high/low confidence triage classes (Methods). To simulate the change in workload over a range of possible prevalences of BE, we first determined the proportion of patients with and without BE in each triage class and then weighted each vector of proportions by a new prevalence ranging from 0 to 55%. To simulate the effect that relative changes in overall sample confidence have on the workload, we first determined the proportion of patients in triage classes with highest sample confidence (determined by quality control and diagnostic class: 2 and 8) and lower sample confidence (1, 3, 4, 5, 6, and 7). We then modified the proportion of high confidence samples and inversely adapted the proportion of lower confidence samples within a range from -25% to 25%.

Over a fine grid of varying disease prevalence and changes in sample confidence, we observed a negative impact of decreasing cohort BE prevalence and a positive impact of sample confidence on the potential workload reduction (Figure 5a). According to this simulation, in a realistic cohort with a BE prevalence of 7%, we would still be able to reduce the pathology workload by 57%. In order to retain the same workload reduction we observed in the validation cohort, the proportion of samples with high confidence in a realistic cohort would need to be increased by 15%.

**Figure 5:**
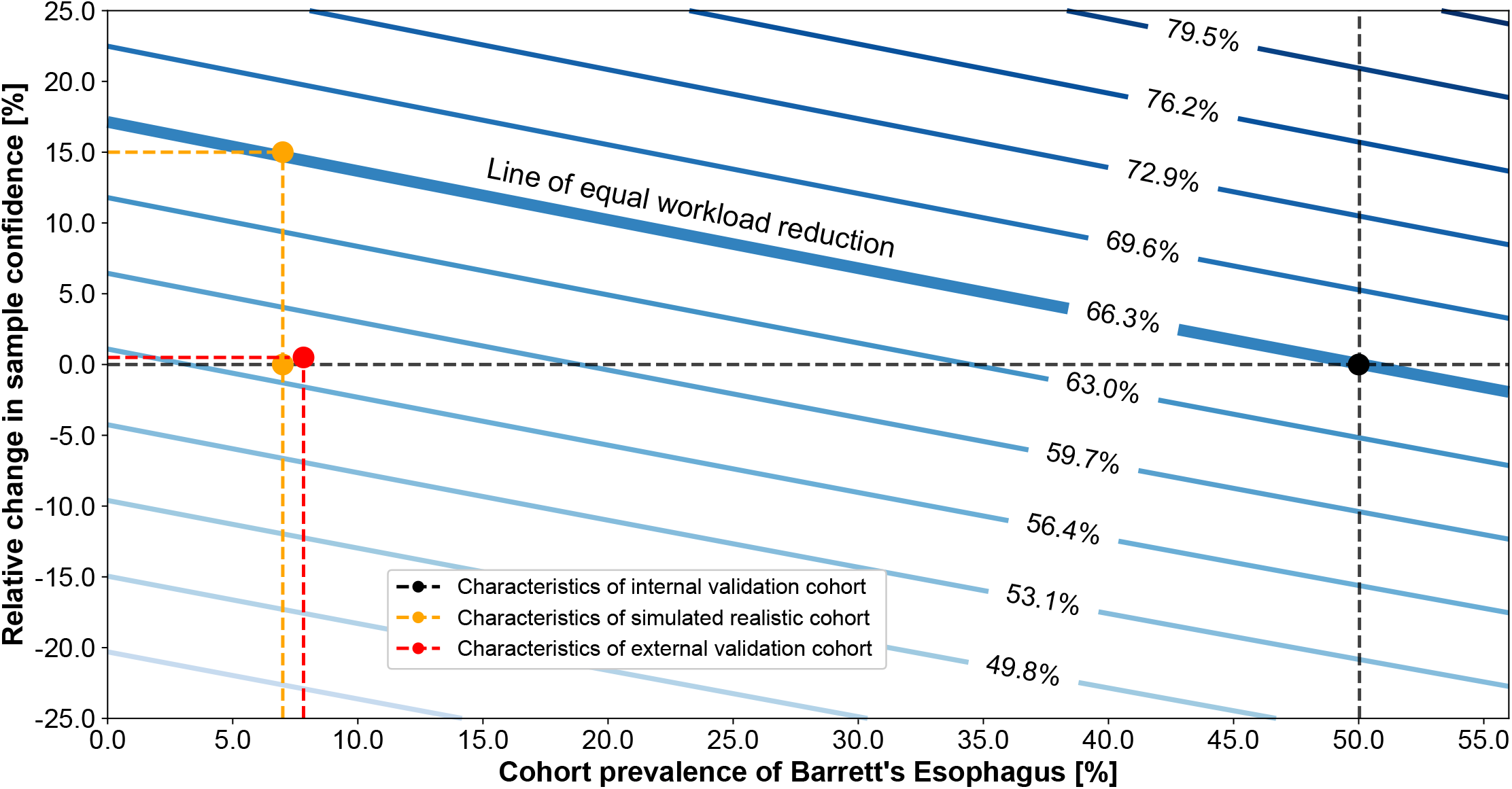
Triage model applied to external validation cohort and simulation of cohort variation. Simulation of changes in cohort prevalence of BE and sample confidence with impact on workload reduction. Every contour line (blue) represents the same level of workload reduction as indicated by the percentages. Solid black lines indicate the workload reduction of the validation cohort. The dotted yellow line illustrates the workload reduction of a realistic primary care referral cohort (with 7% prevalence) with no change in sample confidence classes (lower yellow marker) and the confidence change required to match the same amount of workload reduction as in the validation cohort (upper yellow marker). The results from the external validation cohort are shown in red.

### External validation of triage-driven approach

Finally, we tested the validity of our results and the extrapolation in the simulation study in an independent test set of 3038 slides from 1519 patients from from 109 primary care sites in the UK (BEST3 trial) (*22*). All slides were processed in the same way and with the same model parameters as the BEST2 validation cohort (fig. S7, table S6). Following the method described in the previous section, we used manual pathologist reviews for samples that fell into triage classes 4, 5 and 6. In the BEST3 trial, endoscopy data was only available for positive Cytosponge patients and those who had Barrett’s diagnosed at follow-up as a result of standard of care. In addition, the trial was not designed to investigate sensitivity or specificity but positive predictive value (PPV) instead. We also calculated the negative predictive value (NPV) based on findings aggregated through the primary endpoint analysis (coded BE diagnosis in patient records). For this external validation cohort, fully manual review by pathologists resulted in a PPV of 56.08% and NPV of 99.02%. After application of the triage-driven, semi-automated approach the PPV of the overall cohort was 53.37% and the NPV 99.39% (fig. S7). Using this approach in a realistic primary care setting would have resulted in the following key results: In total 872 patients out of 1519 patients (57.41%) would have been reviewed automatically while 42.59% would have had to be reviewed manually. This agrees with the simulated, expected value (fig. 5) of workload reduction given the prevalence (7.8%) of BE in this external validation cohort. Six additional patients would have been diagnosed with BE while being missed by the pathologist at the cost of 19 additional endoscopies when compared to fully manual review. One patient would have received an automated negative diagnosis even though the pathologist scored it as positive with BE finding at endoscopy.

## Discussion

We have presented a triage-driven approach that analyses samples of the Cytosponge-TFF3 test using deep learning for the early detection of esophageal cancer. Our approach combines quality control and diagnostic metrics of pathology slides to stratify patients into 8 triage classes which determine whether a patient sample requires manual or if automated review would suffice.

For the analysis of Cytosponge-TFF3 samples, our triaging approach has several benefits: We are able to substantially reduce workload and match the sensitivity and specificity of expert pathologists. In our internal validation cohort, fully manual review by a pathologist achieves 81.7% sensitivity and 92.7% specificity. In a fully automated approach, we observed a sensitivity of 72.6% and a specificity of 93.1%. With our triage-driven approach, we demonstrate that up to 66% of cases can be reviewed automatically while achieving a sensitivity of 82.5% and specificity of 92.7%, a performance marginally superior to fully manual review by pathologists. Further, in an external validation cohort from a large randomised controlled trial we observed a PPV of 53.37% and NPV of 99.39%. For comparison, pathologist review resulted in very similar values with a PPV of 56.08% and NPV of 99.02%. While a small number of additional endoscopies would have been triggered, they would have also yielded more positive diagnoses. In this more realistic cohort, 57.41% workload for the pathologists would have been reduced. These results (Figure 5) have several implications: First, a fully automated review would reduce sensitivity (at fixed specificity) and therefore suffer from a loss of clinical utility. Second, while a triage-driven approach is not able to reduce workload as much as a fully automated approach, the described triage classes provide a logical way for stage-wise clinical adoption and performance testing in routine practice.

Another benefit of our approach is that we were able to directly adopt heuristics applied by pathologists familiar with Cytosponge-TFF3 samples in our algorithmic design process. As a result, our approach demonstrates traceability and interpretability (*8*): First, we mimicked the screening process of samples observed by expert pathologists by replicating their decision-making scheme (Figure 1c). Second, the saliency maps we generated from deep learning models to visualize learned features in the pathology images show strong agreement with manual landmarks placed by pathologists (Figure S2).

As a further benefit, our triage approach achieves strong performance from only 287 samples for training and calibration by incorporating informative prior knowledge about biological and clinical test characteristics, followed by rigorous testing in independent cohorts. This performance compares favorably to previous fully automated approaches reporting expert-level performance that relied on very large datasets with training set sizes ranging from 10,000 to more than 100,000 examples (*34, 35*) - dataset sizes that cannot be expected for most applications.

Finally, a quantitative analysis of workload reduction across varying disease prevalences and sample confidences shows that our approach is expected to generalize well to a real-world population. A more general population would have a lower disease prevalence than a case-control study, which would cause a larger workload due to the distribution of BE/non-BE patients within the individual triage classes. We were further able to confirm this simulation with an external validation cohort. These findings provide realistic expectations of how clinical decision-making systems are affected by bias in cohort composition.

Our approach has several limitations: First, while samples used in this work were generated at multiple centres they were processed at only a single site (Addenbrookes Hospital, Cambridge, UK). Thus, our data might not fully reflect the variation in histology sectioning and staining across different laboratories (*36*). We compensated for this limitation through data augmentation by spatial and color profile distortion. Additionally, our data are not too far from future real-world applications, because for large-scale rollout of the Cytosponge test a centralised laboratory is envisaged to ensure processing as well as analysis with proper quality assurance. In future work, we plan to test whether the superiority of the triage-driven approach over fully manual pathologist review will generalize by incorporating multi-centre data from ongoing and future Cytosponge-TFF3 studies to evaluate this effect more extensively.

Second, the underlying machine learning model could be further optimized. For example, instead of using a transfer learning model based on pre-training with a primary dataset, we could train a model from scratch, which has proven to improve results in some CNN applications (*37*). In addition, the tile size needs further investigation because it determines the receptive field in which the CNNs build feature representations of images. Our tile size was chosen by expert pathologists to capture relevant structures like columnar epithelium and goblet cells. Although good performance was observed, a refined multi-scale classification with several magnifications might be necessary to achieve better classification of tissue types. Further improvements might be realised from using attention-based models to reduce the laborious annotation steps required for expanding the training data (*38*) or aggregating tiles to patient level with more sophisticated approaches based on sequence models (*34*).

Third, a major determinant of workload reduction is the quality and therefore diagnostic confidence attributed to a sample. However, what determines the amount of columnar material sampled is unknown. One hypothesis is that the strength of esophageal peristalsis, which can be influenced by variations in device ingestion, may be associated with the likelihood of the Cytosponge reaching the stomach. We plan to investigate determinants of sample quality by comparing the data generated by the trained deep learning models with patient and device administrator profiles.

In summary, our triage approach differs from previous applications of deep learning to medical images (*7,34*) which used fully automated approaches on extremely large datasets. We show that for a modest dataset size, leveraging existing heuristics of pathologist decision-making in a triage-based approach is a powerful alternative to fully automated classification models, which generalises well to an independent validation cohort. These results lay the foundation for tailored, semi-automated decision support systems embedded in clinical workflows.

## Methods

### Study design and dataset

The multicentre Barrett’s Esophagus Screening Trial 2 (BEST2) (*18*) case-control study (study registration: ISRCTN12730505) investigates the automated analysis of Cytosponge-TFF3 samples as a secondary objective. Ethics approval was obtained from the East of England - Cambridge Central Research Ethics Committee (number 10/H0308/71) and registered in the UK Clinical Research Network Study Portfolio (9461). Patients enrolled underwent a Cytosponge procedure followed by an endoscopy with biopsies where required. The objective of this work was the comparison of: fully manual review of Cytosponge-TFF3 pathology slides by human experts, fully automated review of Cytosponge-TFF3 pathology slides by a deep learning-based method, and triage-driven, semi-automated review of Cytosponge-TFF3 pathology by a hybrid method relying on deep learning methods and human experts.

812 patients were randomly selected from the entire BEST2 cohort (from 11 hospitals in the UK) for digitisation of their respective H&E and TFF3 pathology slides (1624 in total) on an Aperio AT2 digital whole-slide scanner (Leica Biosystems Nussloch GmbH, Germany) at 40x magnification.

BEST2 patients were randomly partitioned into three distinct subsets: 100 patients for training/development (labels unblinded for training purposes), 187 patients for calibration (labels unblinded for calibration), and 525 patients as an internal validation set (labels unblinded after validation). The distribution of patients with or without Barrett’s Esophagus (BE) for each partition is shown in Figure 1d.

For independent external validation we used data from the Barrett’s Esophagus Screening Trial 3 (BEST3) REF randomised controlled trial (study registration: ISRCTN68382401). Ethics approval was obtained from the East of England - Cambridge Central Research Ethics Committee (number 16/EE/0546). Patients enrolled either were invited to a Cytosponge procedure or received standard of care. Both arms were followed up after 8 to 18 months (weighted overall average of approx. 12 months). Only patients who underwent a Cytosponge procedures or were referred as part of usual care received an endoscopy. A patient was considered as positive for Barrett’s Oesophagus if they either had a diagnosis at endoscopy or as a result of a coded search in records from the primary care site.

1519 patients were randomly selected from the entire BEST3 cohort (from 109 primary care sites in the UK) for digitisation of their respective H&E and TFF3 pathology slides (1638 in total) on Hamamatsu S60 and S210 whole-slide scanners (Hamamatsu, Japan) at 40x magnification. For each patient, the repeat test was used if one as performed due to inadquace of the baseline test.

All BEST3 patients were processed using the fully automated and triage-driven, semi-automated approach presented in this work. Labels were unblinded after validation.

Confidence intervals in this work were defined as the 2.5th and 97.5th percentiles on distributions of 500 samples (with replacement) of the respective dataset size.

### Cytosponge-TFF3 procedure

The Cytosponge-TFF3 is a non-endoscopic diagnostic modality for BE. It is a cell collection device, consisting of a mesh sphere on a string inside a gelatine capsule, coupled with an immunohistochemical biomarker called Trefoil Factor 3 (TFF3).

The capsule is swallowed by the patient, and passes to the stomach, where the gelatine dissolves allowing the mesh sphere to expand to a diameter of 3 cm. After 5 to 7.5 minutes, the sponge is withdrawn from the stomach by the attached string, sampling superficial epithelial cells from the top of the stomach, the esophagus, and the oropharynx. The removed device is placed in a container with preservative solution (SurePath Preservative Fluid, BD) and processed in a laboratory for histochemical (Hematoxylin & Eosin) and immunohistochemical (TFF3) staining. The stained pathology slides are then screened by a pathologist. The primary objective of the Cytosponge-TFF3 test is the detection of columnar epithelium of intestinal type (with TFF3-positive goblet cells) in the squamous oesophagus which is indicative of the patient having Barrett’s Esophagus (BE). These TFF3-positive patients can then be referred for an upper gastrointestinal endoscopy to confirm the diagnosis. Previous studies (*17–19*) have shown a consistent sensitivity (73.3 % and 79.9 %) and specificity (93.8 % and 92.4 %) for the diagnosis of BE using the Cytosponge coupled with TFF3.

### Endoscopy procedure

Esophago-gastroduodenoscopies were carried out by an endoscopist after the Cytosponge test. BE was defined as endoscopically visible columnar-lined esophagus that measured at least 1 cm circumferentially or at least 3 cm in non-circumferential tongues according to the Prague criteria (≥C1 or ≥M3 (*39*)). An additional criterion for BE was histopathological evidence of intestinal metaplasia (IM) on at least one endoscopy biopsy. For cases with suspected BE, diagnostic biopsies were collected following the recommended Seattle surveillance protocol (*40*). When reviewing the biopsy data, all of the pathologists were blinded to the result of the Cytosponge-TFF3 test.

### Whole-slide image annotation for training

One H&E- and one TFF3-stained slide for each of the 100 BEST2 patients from the training set were manually annotated and reviewed by an expert pathologist (MO) using the ASAP software (*41*). Regions of interest (ROIs) were selected in the digitised pathology slides at a magnification of 40x. Each of these ROIs was labeled with a class for training. For the H&E-based quality control model, four different classes were identified: gastric-type columnar epithelium, respiratory-type columnar epithelium, intestinal metaplasia, and background (including other cellular material such as squamous cells and slide artefacts). Gastric-type columnar epithelial cells were considered as the marker for quality control, as their presence confirms that the Cytosponge has reached the stomach. For the TFF3-based diagnostic model, three classes were identified: TFF3-positive regions (darkly stained goblet cells), TFF3-equivocal regions (regions of ambiguous staining that may be goblet cells), and background. TFF3-positive cells were considered as the marker for the presence of IM, as they indicate that the patient might have BE. All slides were annotated using the existing patient-level ground truth data for comparison. We aimed for a representative fraction of available material on each slide to be labelled.

### Tesselation of whole-slide images for training

Tesselation, or tiling, of whole-slide images was performed in order to prepare data prior to model training. A custom tiling method was developed to optimise the yield and coverage of annotated cellular material in the images. Whereas packing problems of squares in polygons can be neglected for large annotations, optimal coverage for tiles in combination with small annotation sizes is not straightforward and requires a tailored solution. Annotations with an area of 1.5 ∗ *tile area* or larger were cropped into tiles by taking the top-left coordinate of the enveloping bounding box and iterating tiles along the x- and y-axis of the image. Tiles with an intersection of less than 0.33 (for H&E) or 0.66 (for TFF3) with their corresponding annotation were rejected. Annotations with an area smaller than 1.5 ∗ *tile area* were treated as single examples and a tile was placed in the center-of-mass of the respective annotation. Tiles with sufficient annotation coverage (determined by intersection) were extracted and labelled according to the class of their parent annotation. For this work, a tile size of 400-by-400 pixels (corresponding to 200-by-200 µm at a magnification of 40x) was selected in accordance with sizes of relevant tissue features. Tiles were extracted from whole-slide images as JPEG images with minimal compression.

### Model training using deep learning

We implemented two different deep learning frameworks: one for performing quality control on H&E-stained slides, and a second one for performing automated BE diagnosis from the TFF3-stained slide images. Both deep learning frameworks for quality control and diagnosis were created by comparative transfer learning of multiple convolutional neural network architectures: AlexNet (*23*), DenseNet (*24*), Inception v3 (*25*), ResNet-18 (*26*), SqueezeNet (*27*), and VGG-16 (*28*). All architectures were initialised with the best parameter set that was achieved on the ImageNet competition. Training tile images were resized as required for the individual architectures, resulting in a change of effective magnification from 22x to 30x. We then unfroze all layers to enable fine-tuning of the entire network. For all models, training continued on two NVIDIA GTX 1080Ti graphics cards for 25 epochs with an architecture-specific batch size (ResNet-18: 128, VGG-16: 48, Inception v3: 48, AlexNet: 64, SqueezeNet: 256, DenseNet: 84) and a learning rate that decayed by a factor of 0.1 every 7 epochs. All models used cross-entropy loss. To account for slight variations in the training data, random vertical/horizontal flip, random rotation, and random color jitter (variation in hue, contrast, brightness, and saturation) were introduced for data augmentation. Differences in tile class sizes were accounted for by using a modified imbalanced dataset sampler, a function which oversamples from minority classes and undersamples from majority classes. The parameter set of epoch with the highest accuracy on the development subset was selected for further use. All models were trained using the PyTorch deep learning framework (*42*). Final model versions used a split of 85:15 patients for training and development subset. We further investigated the effect of increased training set sizes by incrementally increasing the training subset while fixing the development subset size (Figure S1).

### Evaluation of tile-level performance

In order to compare the performance of all six deep learning architectures, we calculated class-specific performance in the quality control and diagnosis frameworks (Table S1). To obtain these numbers, we selected the epochs with the best weighted accuracy score on the development subset for each training run. We then calculated precision and recall of all four classes in the H&E-based model and all three classes in the TFF3-based model in the selected epoch. For visual comparison, we also created 2D inference maps of samples which where classified as positive or negative by a pathologist for quality control and diagnosis, respectively. Tile-level results were not used to select architectures for the fully automated or semi-automated, triage-driven approach. The best performing architectures according to relevant class precision and recall on tile level for quality control and diagnosis were selected for saliency map generation.

### Generation of saliency maps using Grad-CAM

Gradient-weighted Class Activation Mapping (Grad-CAM) class localisation maps are created by visualising the gradients flowing into the final convolutional layer of the network, just before the fully-connected layers (*29*). Since convolutional layers contain class-specific spatial information from the input image which is lost in the fully connected layers, this is the optimal point for map generation. Unlike conventional class-activation maps (CAMs), Grad-CAM has the benefit of not requiring any modifications to the existing model architecture, nor does it require any retraining of the model (*29*). In order to create the class-specific Grad-CAM localisation map for class *c*, 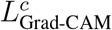, it is first necessary to compute the gradient 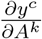 of the score *y*^*c*^ for class *c* with respect to the feature map *A*^*k*^ of the final convolutional layer (*29*). Once 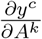 has been computed for each feature map *k*, these backward-flowing gradients are global-average-pooled across the width and height of the network (indexed by *i* and *j*) to yield 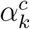, the weights of neuron importance for each of the feature maps *k* (*29*):

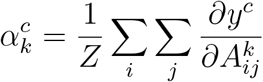

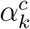, the neuron-importance weights for each feature map *k*, therefore estimate the salience of each feature map to the prediction of class *c* (*29*). Finally, to get class *c*-specific Grad-CAM localisation map 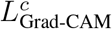, we take the *ReLU* of the weighted sum of the feature maps *A*^*k*^, where each feature map *k*’s weight is 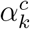 (*29*):

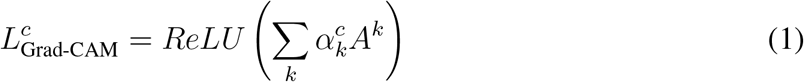

Note that the *ReLU* operation is used to retain only the features which have a positive influence on the prediction of class *c*, and that the resulting localisation map will be the same size as the feature maps of the last convolutional layer (*29*).

We generated saliency maps for both models trained on H&E and TFF3, respectively. The target layer from the VGG-16 architecture was the last feature layer (no. 30) before several stacked fully connected layers. Tiles were randomly selected from the development subset. For qualitative comparison between saliency maps and manual landmarks, we asked one expert pathologist (MO) to highlight important areas. Areas highlighted by the pathologist provide a representation of features which a human observer uses for classification of tile images. To investigate qualitative agreement of landmarks by the pathologist with generated saliency maps, a side-by-side comparison of tile images and respective saliency maps was prepared (Figure S2).

### Model inference on calibration and validation cohort

All six deep learning architectures trained separately for quality control and diagnosis tasks were applied to pathology slides randomised to calibration and validation cohort. Whole-slide images were tesselated on the fly as described above. Detection of tissue was achieved by luminance thresholding of tile values in the LAB colour space. Tiles were forward-passed through the trained deep learning architectures and softmax probabilities were aggregated for each tile position.

### Aggregation of classifications on tile level to the patient level

We explored two different aggregation approaches based on propagation of the individual tile-level classifications to patient-level classifications for quality control and diagnosis: a fully automated approach which operates on the basis of a single operating point, and a semi-automated, triage-driven approach which leverages two operating points. For the former approach, performance was assessed using sensitivity and specificity; for the latter, performance was assessed using an incremental substitution scheme with simultaneous analysis of sensitivity and specificity. For both approaches, tile-level probabilities had to be thresholded to obtain the number of positive tiles per slide for quality control and diagnosis. In the following section, we describe how tile-level probabilities were thresholded and how the operating points on the resulting numbers of positive tiles (quality control and diagnosis) were then calibrated and evaluated as part of each approach.

### Determination of tile-level probability thresholds

In order to generalise the tile-level probabilities to the number of positive tiles per patient, we determined thresholds for each model and endpoint (quality control and diagnosis). The probability threshold of individual tiles for quality control and diagnosis had to be determined, then, the resulting number of positive tiles per threshold was assessed against the best ROC-AUC on the calibration cohort (Figure S3, Table S2).

To achieve the best-performing threshold for individual tile probabilities and subsequent aggregation, we iterated over a range of tile thresholds on a fine grid from 0 to 1 (in 0.005 steps and inclusive of 0.999, 0.9999, and 0.99999). For the quality control model on H&E, the relevant class was gastric-type columnar epithelium. For the diagnosis model on TFF3, the relevant class was TFF3-positive goblet cells.

In order to determine the resulting number of positive tiles per threshold, probability thresholds for quality control were compared (ROC-AUC) to the pathologist ground truth of H&E slide analysis. Probability thresholds for diagnosis were compared (ROC-AUC) to endoscopy (confirmation of BE presence by endoscopist and IM on endoscopy biopsy by pathologist) ground truth. This step was required to determine the optimal threshold for individual tile classification. This threshold was then used in the calibration and validation of the fully automated and semi-automated, triage-driven model as described in the next section.

### Calibration of fully automated model

All six deep learning architectures trained for quality control and diagnosis were applied to the whole-slide images from the calibration cohort (see Model inference). The number of positive tiles per sample for quality control and diagnosis was determined as described above. To determine an adequate operating point for the fully automated patient-level model, ROC analysis was performed on the number of detected tiles (quality control and diagnosis) per patient. On the same set of patients, we calculated the performance by an expert pathologist. In order to determine the ideal cut-off for number of detected tiles, we fixed the specificity of each model to the performance of an expert pathologist on the calibration cohort. The resulting operating point was then chosen for validation of the fully automated model in the validation cohort (Table S2). Tile-level thresholds which yielded the best sensitivity on the calibration cohort were used for evaluating all approaches on the validation cohort. The best-performing architecture (assessed by sensitivity) on the calibration cohort was considered the representative model for application on the validation cohort. However, due to the simplicity of operating point determination, the performance of all other architectures on the validation cohort was also investigated.

### Evaluation of fully automated model using ROC analysis

All six deep learning architectures trained for quality control and diagnosis were applied to the whole-slide images from the validation cohort (see Model inference). The number of positive tiles per sample for quality control and diagnosis was determined as described above. Subsequently, the previously determined operating point (calibration) for each of the deep learning architectures was applied. The binary results were then compared against ground truth of the quality control and diagnosis models. For quality control on H&E, the results were compared to the ground truth of the pathologist who was reading the H&E slide of the Cytosponge test. For diagnosis on TFF3, the results were compared with endoscopy ground truth (with confirmation of BE presence by endoscopist and IM on endoscopy biopsy by pathologist). Sensitivities and specificities on the validation cohort were calculated for all models with an additional presentation of ROCs for visualisation (Table S3, Figure S5). For comparison with other approaches, performance metrics of the architecture selected during calibration of the fully automated model were used.

### Calibration of triage-driven, semi-automated model

All six deep learning architectures trained for quality control and diagnosis were applied to the whole-slide images from the calibration cohort (see Model inference). For calibration, only the best model (according to ROC-AUC) was presented to two expert observers to determine operating points. The number of positive tiles per sample for quality control and diagnosis was determined as described above (Figure S6). The objective of this approach was a more granular classification of patients into three classes for quality control and diagnosis and subsequent stratification by different class combinations. Therefore, two operating points were determined for each model, instead of one.

Both observers were presented with the number of detected tiles and relevant ground truth (Cytosponge pathology and endoscopy) for quality control and diagnosis models. They were instructed to choose two operating points for each task: First, an operating point which optimises sensitivity with a low number of false positives. Second, an operating point which separates the intermediate region of the first and second operating point from samples with optimised specificity and a low number of false negatives. The resulting operating points were then chosen for validation of the semi-automated, triage-driven model in the validation cohort (Table S5).

The two operating points for quality control and diagnosis resulted in three tiers per framework and were labelled as follows: for quality control, samples above the first operating point were to be considered as high confidence, samples between the first and second operating point as low confidence, and samples below the second operating point as no confidence. For diagnosis, samples above the first operating point were to be considered as high confidence positive, samples between the first and second operating points as low confidence equivocal, and samples below the second operating point as high confidence negative. Eight triage classes (number 1 to 8) were composed by all possible combinations of quality control and diagnosis classes. The combination (no confidence in quality and high confidence in diagnosis) is likely artifactual and was therefore merged (with no confidence in quality and equivocal in diagnosis) to form triage class 4. Two expert observers then ranked all eight classes from lowest to highest likelihood for patients having BE. They further assigned a qualitative rank for priority of manual review based on the subjective difficulty to review samples that are part of specific triage classes.

### Evaluation of triage-driven model on internal validation cohort

The triage-driven, semi-automated model was evaluated by applying a cumulative substitution scheme on the internal validation cohort. The base scenario for all cumulative substitutions was the performance of the pathologists on the entire validation cohort. At every substitution, the pathologists’ Cytosponge-TFF3 results were substituted with automated review in the respective triage classes. Then, sensitivity, specificity, and proportion of patients substituted with automated review were calculated and compared against the previous substitution steps. The substitution scheme was applied starting from both ends of the triage class list. First, class 1 was substituted with automated review, then classes 1 and 2, then classes 1, 2, and 3, and so on. Second, class 8 was substituted with automated review, then classes 8 and 7, then classes 8, 7, and 6, and so on. We then analysed the sensitivity and specificity curves for deviations from their previous values for each step in both applications of the scheme. Classes which caused a drop in sensitivity or specificity on substitution were considered as ‘difficult’ and we retained manual review by a pathologist for associated samples. For each of the difficult classes we then summed up the number of patients that fell into these classes and divided by the total number of patient in the validation cohort. This ratio was to be considered as the potential workload reduction which this substitution scheme could achieve without notable loss in performance.

### Simulation of cohort variation and impact on workload reduction

In order to assess workload reduction in cohorts with different compositions, we simulated the distribution of patients within triage classes with varying BE prevalences and sample confidences. Let *P* be a set of all patients with two subsets: *Q* ⊆ *P* contains all patients with BE and its complement *R* = *P \ Q* contains all patients without BE. We count the proportions of patients in each triage class in each of the sets *P, Q, R* as vectors **c**^*P*^, **c**^*Q*^ and **c**^*R*^, respectively. Our simulation consists in re-weighting these vectors to reflect different BE prevalences and sample confidences. For each element of a range of BE prevalences (**s**_prev_ = {0.00, 0.01, …, 0.55}) we multiply **c**^*Q*^ by *s ∈* **s**_prev_ and **c**_*R*_ by 1 − *s*. At the same time, for each element of a range of relative sample confidences (**t**_conf_ = {−0.25, −0.24, …, 0.25}) we shift proportions of **c**^*P*^ between triage classes {1, 3, 4, 5, 6, 7} and {2, 8} by adding *t ∈* **t**_conf_ to one set of classes and subtracting it from the other. Reduction of workload (*W*) at every simulation step was defined as **c**^*P*^ for classes 4, 5, and 6 over classes 1, 2, 3, 7, and 8:

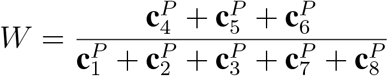

### Evaluation of triage-driven model on external validation cohort

The triage-driven, semi-automated model was further evaluated applying it with frozen model parameters on the external validation cohort. Processing of images was performed as described on the internal validation cohort above. The trial from the data originates was investigating real-world implementation of the Cytosponge device technology. Therefore, endoscopy data endoscopy data was only available for positive Cytosponge patients and those who had Barrett’s diagnosed at follow-up as a result of standard of care. This resulted in a difference of available data as the study was designed for PPV instead of sensitivity and specificity. The NPV was also calculated by using aggregated findings from the primary trial endpoint. An analysis according to the presented substitution scheme was additionally performed ()

### Code availability

The source code of this work is freely available at a public repository: https://github.com/markowetzlab/cytosponge-triage.

### Data availability

The dataset is governed by data usage policies specified by the data controller (University of Cambridge, Cancer Research UK). We are committed to complying with Cancer Research UK’s Data Sharing and Preservation Policy. Whole-slide images used in this study will be available for non-commercial research purposes upon approval by a Data Access Committee due to institutional requirements. Applications for data access should be directed to. Data derived from the raw images are freely available at a public repository: https://github.com/markowetzlab/cytosponge-triage. The code and included data enable replication of the results and figures in this manuscript.

## Data Availability

The dataset is governed by data usage policies specified by the data controller (University of Cambridge, Cancer Research UK). We are committed to complying with Cancer Research UK's Data Sharing and Preservation Policy. Whole-slide images used in this study will be available for non-commercial research purposes upon approval by a Data Access Committee due to institutional requirements. Applications for data access should be directed to.
Data derived from the raw images are freely available at a public repository: https://github.com/markowetzlab/cytosponge-triage. The code and included data enable replication of the results and figures in this manuscript.

## Acknowledgments

This research has been supported by Cancer Research UK (FM: C14303/A17197), the Medical Research Council (RCF: RG84369) and Cambridge University Hospitals NHS Foundation Trust. BEST2 was funded by Cancer Research UK (12088 and 16893). MG acknowledges support from an Enrichment Fellowship from the Alan Turing Institute. MCO acknowledges support from a Borysiewicz Fellowship from the University of Cambridge and Junior Research Fellowship from Trinity College, Cambridge. FM is a Royal Society Wolfson Research Merit Award holder. We thank Michael Schneider, Ruben Drews, Paula Martinez-Gonzalez, and Tristan Whitmarsh for valuable input on this work. The authors thank the Cambridge Biomedical Research Centre and the Experimental Cancer Medicine Centre for their support and for providing the infrastructure for the research procedures in Cambridge. Further, we thank the Human Research Tissue Bank, which is supported by the UK National Institute for Health Research (NIHR) Cambridge Biomedical Research Centre, from Addenbrookes Hospital. Last, we thank the BEST2 trial team, the Histopathology core facility at the Cancer Research UK Cambridge Institute, and Pathognomics Ltd for their support.

## Author contributions

MG conceived and led the analysis; MCO and AB contributed to the analysis; MG and AB wrote the code for analysis; MO and RCF were involved in collection and labelling of the data; RCF conceived the study; RCF and FM directed the project; MG and FM wrote the manuscript with the assistance and feedback of all other co-authors.

## Supplementary materials

### Figures

**Figure S1:**
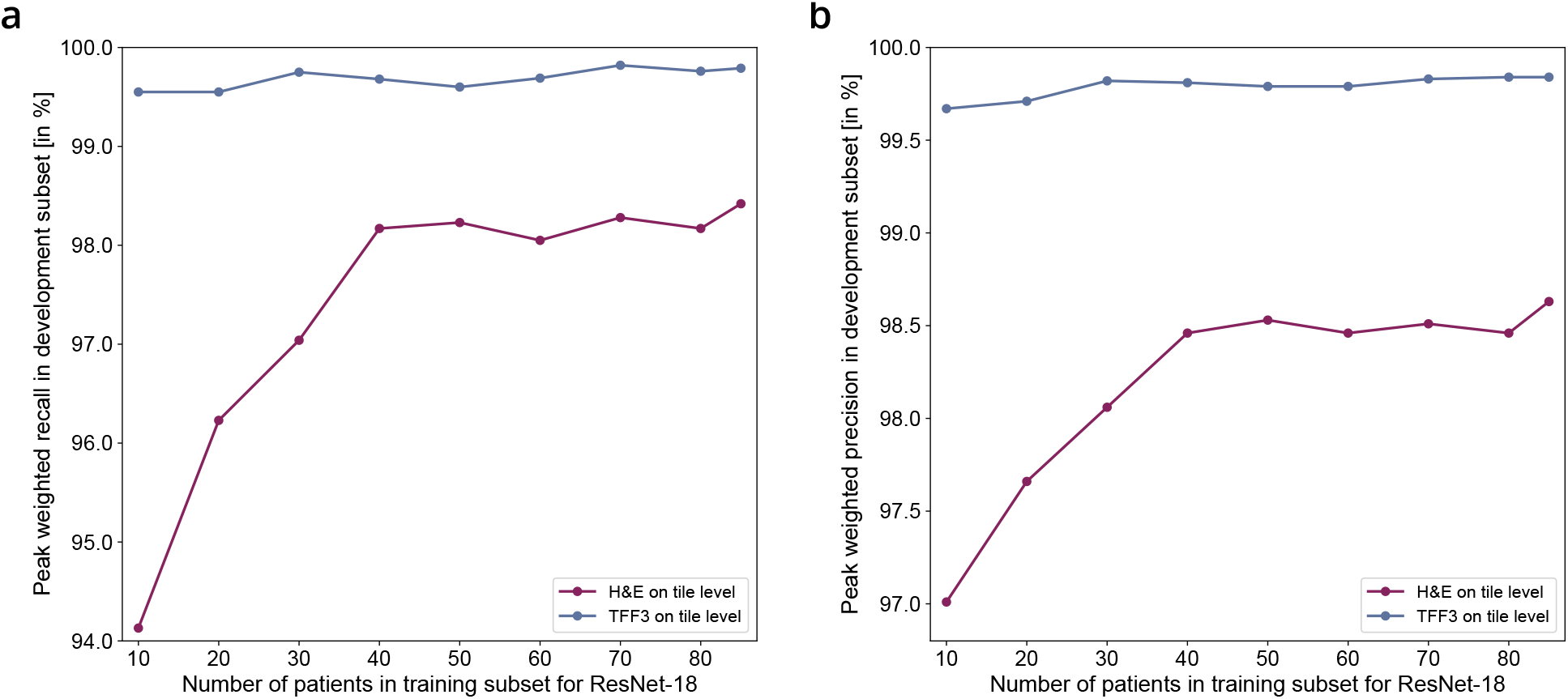
Differential increase of training partition size for ResNet-18. Training subset refers to the relative proportion of the training partition used in the model training phase. Development subset refers to the relative proportion of the training partition used in the model development phase. The peak development weighted recall (a) and precision (b) correspond to the best performing cohort for each training run. The size of the development set was fixed at 15 patients. For each patient, an average of 3,500 tiles was used. For both H&E and TFF3 no substantial increase in performance metrics could be observed after a training subset size of 50 patients. H&E benefited more from an increased number of patients than the TFF3 model. This difference is associated with the increased complexity of detecting different tissue morphologies on H&E vs. brown goblet cells on TFF3.

**Figure S2:**
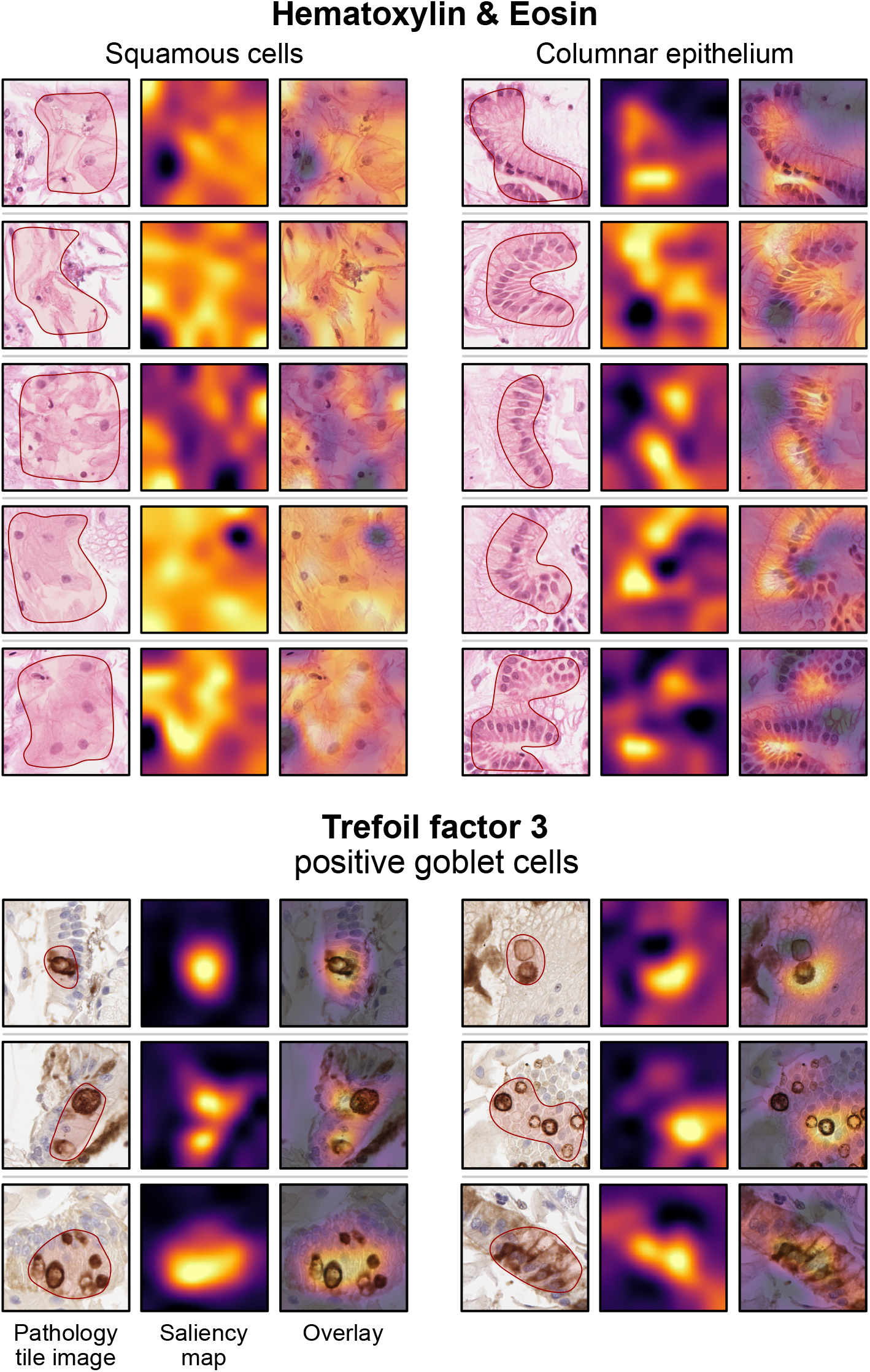
Comparison of pathologist landmarks with saliency maps extracted from VGG-16 architectures. Additional examples of saliency maps for Hematoxylin & Eosin stain (squamous cells and columnar epithelium) and Trefoil factor 3 (positive goblet cells). Landmarks selected by an expert pathologist are shown as overlays with red borders on pathology tile images. For all classes, there was visual agreement between highlighted areas by the pathologist and saliency map activations.

**Figure S3:**
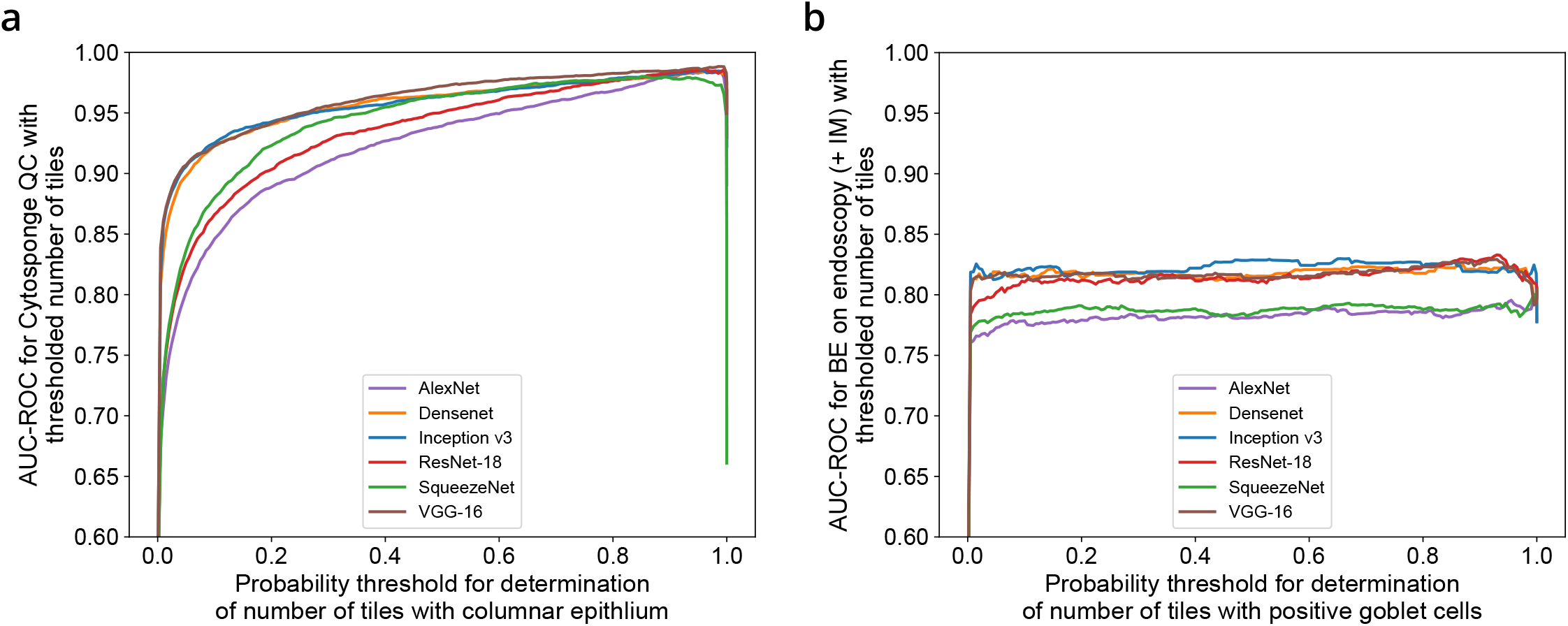
Determination of probability thresholds in order to obtain number of tiles. Both plots show the AUC-ROC for individual probability thresholds (after softmax) which are used to decide whether a tile falls into the relevant class. (a) AUC-ROC for quality control (QC) ground truth determined by the pathologist compared with number of tiles containing columnar epithelium at individual probability thresholds. (b) AUC-ROC for diagnosis ground truth determined by the endoscopy (with confirmed IM on pathology) compared with number of tiles containing positive goblet cells at individual probability thresholds.

**Figure S4:**
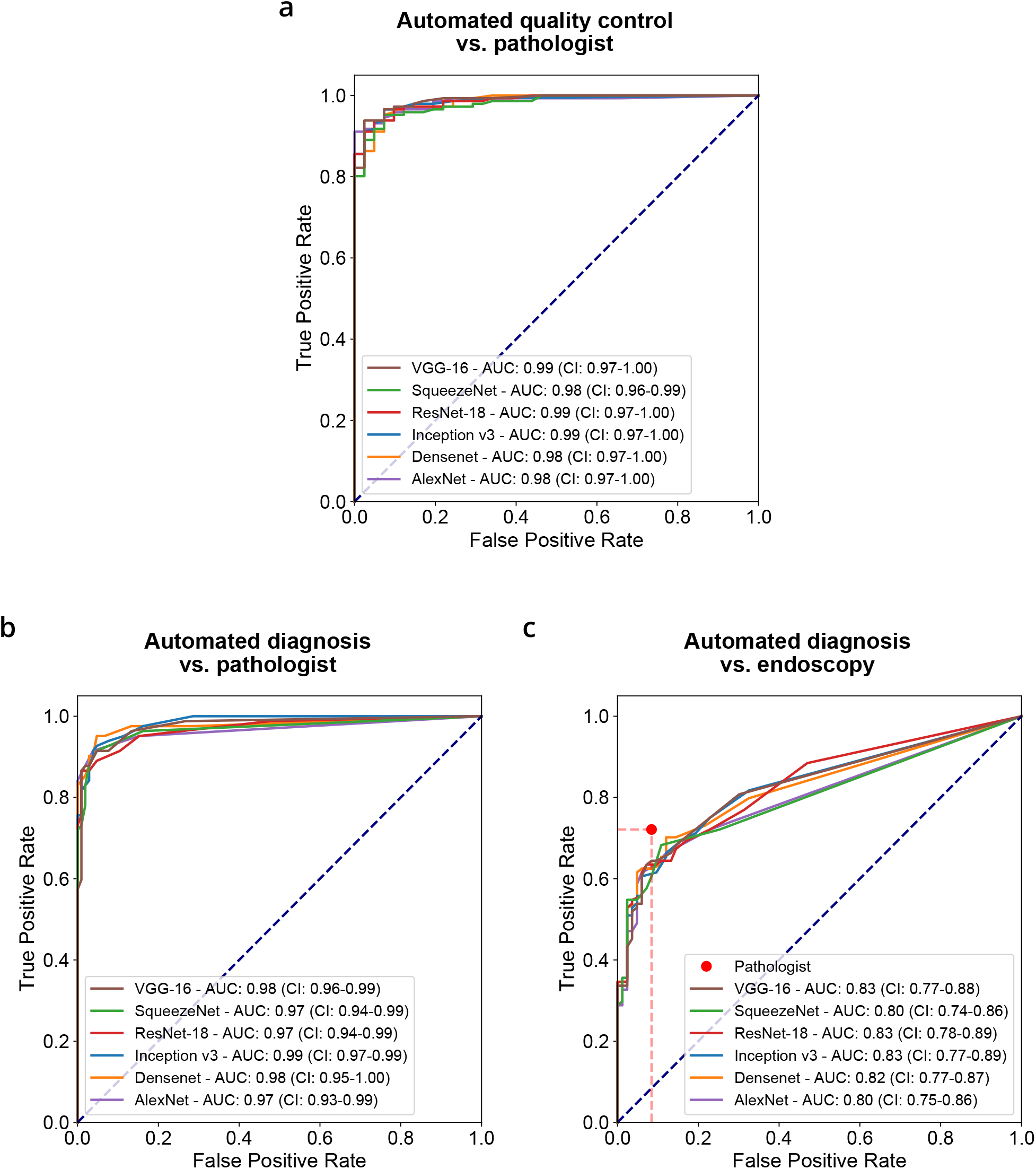
Performance of all deep learning architectures on the calibration cohort. (a) ROC analysis of number of tiles containing columnnar epithelium on H&E compared with pathologist ground truth from Cytosponge (b) ROC analysis of number of tiles containing positive goblet cells on TFF3 compared with pathologist ground truth from Cytosponge (c) ROC analysis of number of tiles containing positive goblet cells on TFF3 compared with endoscopy (with confirmed IM) ground truth. A weak AUC dependency on architecture complexity can be observed.

**Figure S5:**
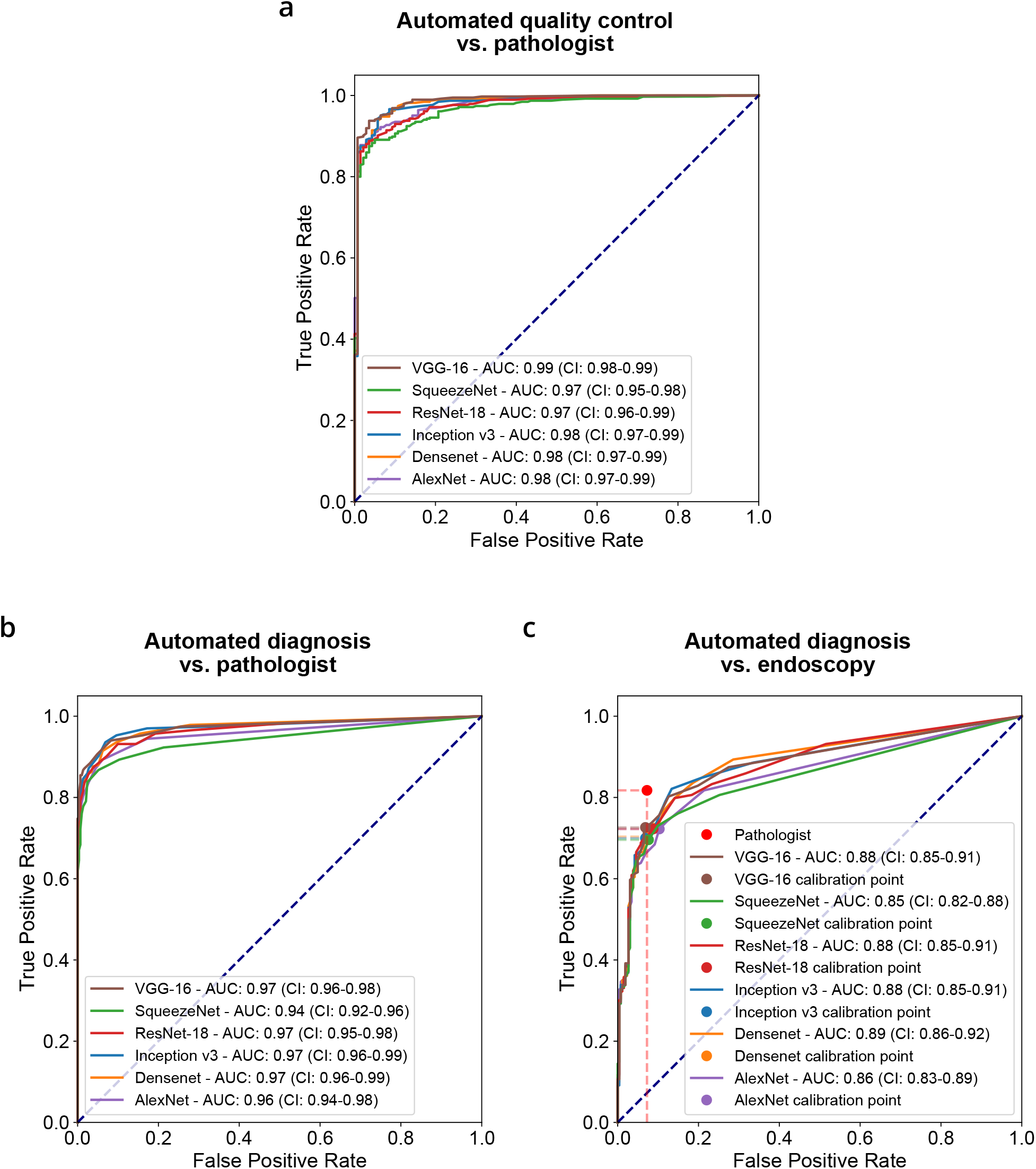
Performance of all deep learning architectures on the validation cohort. (a) ROC analysis of number of tiles containing columnnar epithelium on H&E compared with pathologist ground truth from Cytosponge (b) ROC analysis of number of tiles containing positive goblet cells on TFF3 compared with pathologist ground truth from Cytosponge (c) ROC analysis of number of tiles containing positive goblet cells on TFF3 compared with endoscopy (with confirmed IM) ground truth. As in the calibration cohort, a weak AUC dependency on architecture complexity can be observed.

**Figure S6:**
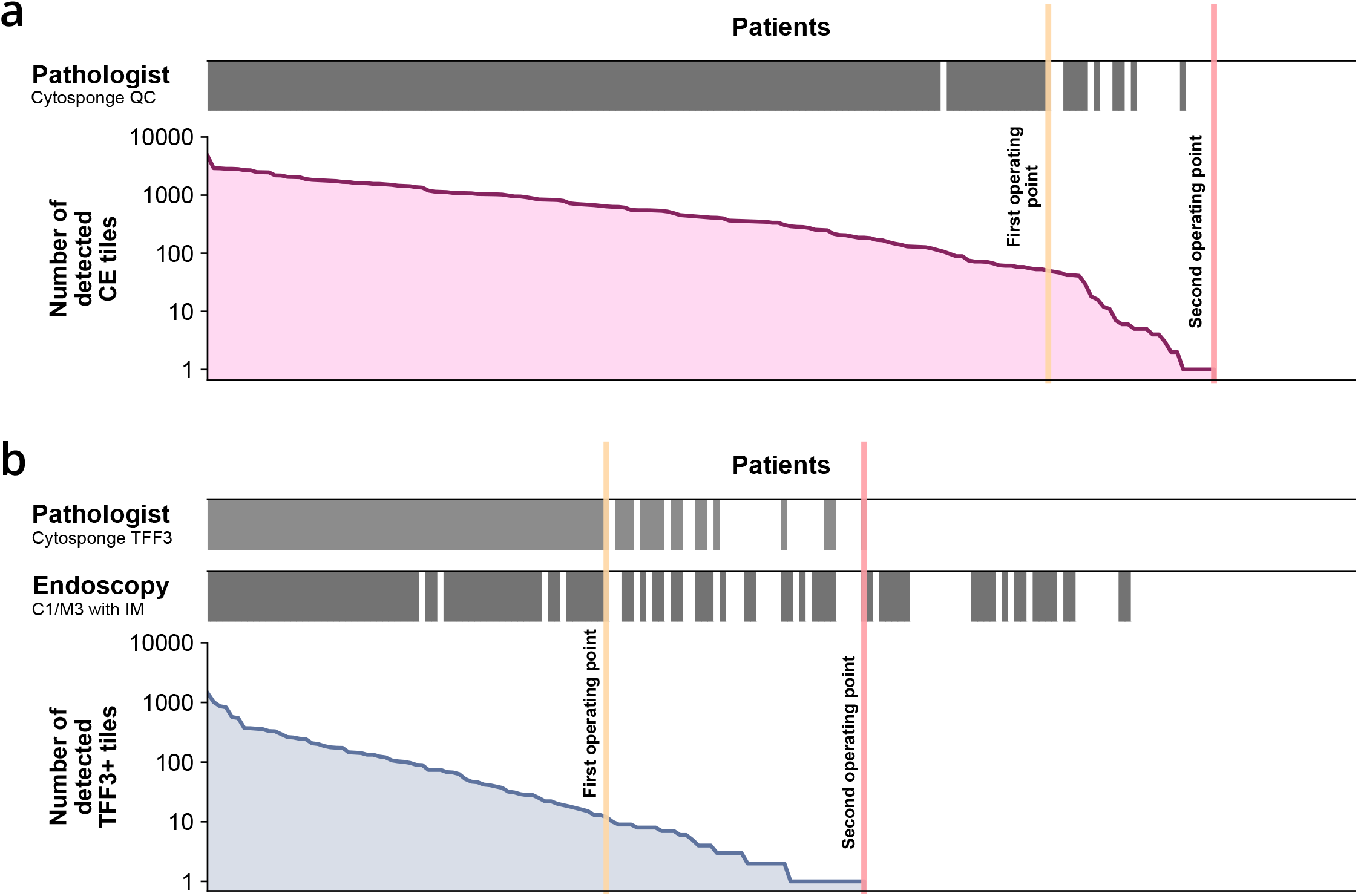
Application of quality control and diagnostic confidence class scheme to calibration cohort. **a** Quality ground truth by pathologist from Cytosponge (top) compared with number of detected columnar epithelium (CE) tiles on H&E detected by VGG-16 (bottom). **b** Diagnosis ground truth by pathologist from Cytosponge (top), Endoscopy (with confirmed IM on biopsy) ground truth (middle) compared with number of detected TFF3-positive tiles on TFF3 detected by ResNet-18 (bottom) / eqv. = equivocal.

**Figure S7:**
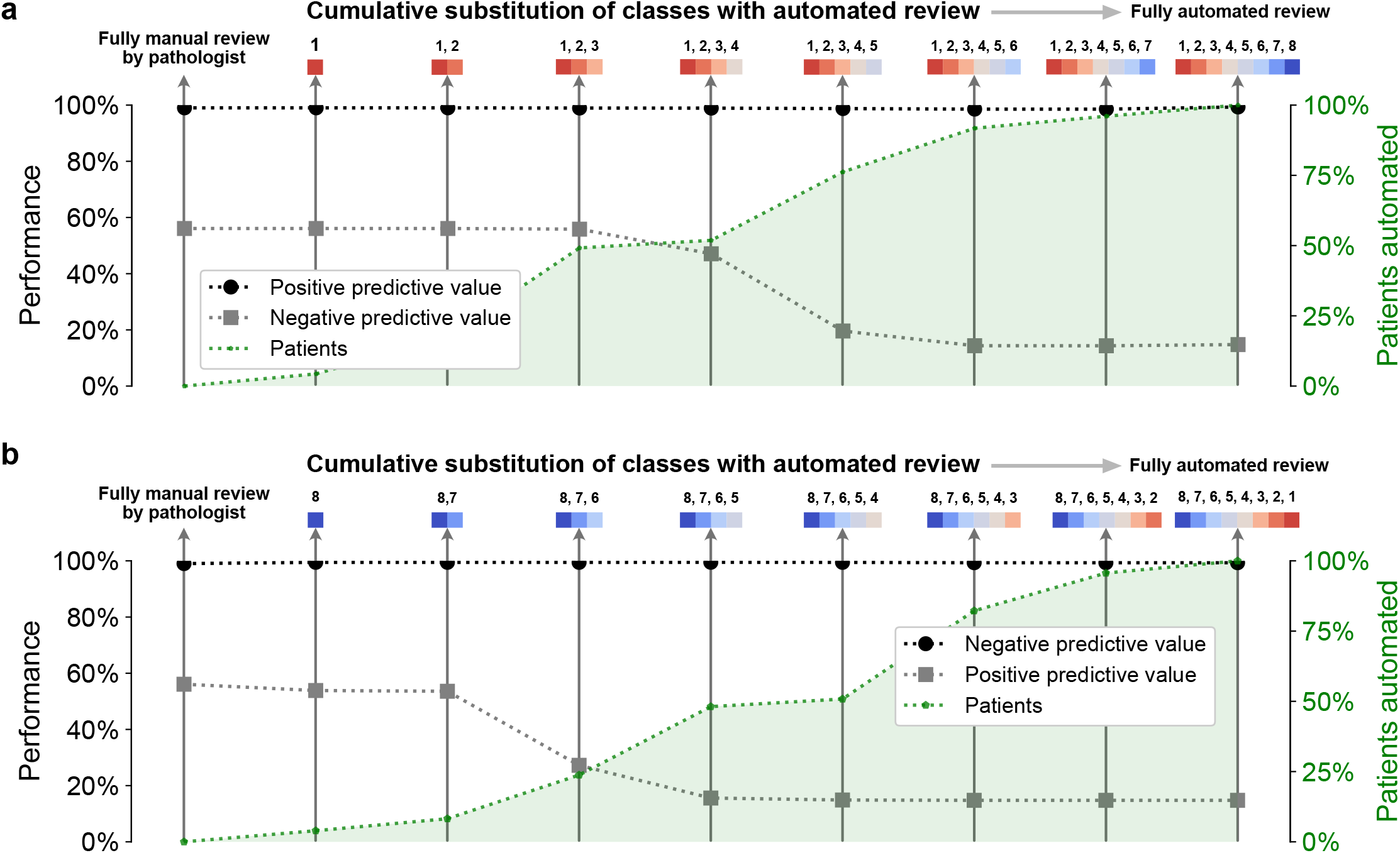
Performance of semi-automated, triage-driven model on external validation cohort. **a** Cumulative substitution scheme starting with fully manual review, followed by substitution with automated review of class no. 1, then 1 and 2, etc. **b** Cumulative substitution scheme starting with fully manual review, followed by substitution with automated review of class no. 8, then 8 and 7, etc.

### Tables

**Table S1:** Tile-level precision and recall for all classes from H&E and TFF3 models. This data is derived from the tiles in the development set. (DenseNet = DenseNet-121, Inception = Inception v3, ResNet = ResNet-18, VGG = VGG-16). The highest value(s) per row is/are highlighted in bold.

|  | AlexNet | DenseNet | Inception | ResNet | SqueezeNet | VGG |
| --- | --- | --- | --- | --- | --- | --- |
| **H&E** |  |  |  |  |  |  |
| Overall accuracy | 0.977 | **0.990** | 0.989 | 0.984 | 0.959 | 0.988 |
| **Precision** |  |  |  |  |  |  |
| Background | 0.999 | 0.999 | 0.999 | 0.999 | 0.999 | 0.999 |
| CE (gastric type) | 0.791 | **0.865** | 0.857 | 0.807 | 0.763 | 0.843 |
| CE (respiratory type) | 0.389 | 0.750 | **0.895** | 0.667 | 0.241 | 0.741 |
| Intestinal Metaplasia | 0.393 | **0.688** | 0.609 | 0.518 | 0.215 | 0.640 |
| **Recall** |  |  |  |  |  |  |
| Background | 0.984 | 0.995 | **0.996** | 0.991 | 0.963 | 0.995 |
| CE (gastric type) | 0.893 | 0.947 | 0.940 | 0.921 | 0.935 | **0.950** |
| CE (respiratory type) | 0.802 | 0.779 | 0.588 | 0.794 | **0.832** | 0.634 |
| Intestinal Metaplasia | 0.606 | 0.610 | 0.629 | 0.606 | **0.643** | 0.568 |
| **TFF3** |  |  |  |  |  |  |
| Overall accuracy | 0.996 | **0.999** | 0.998 | 0.998 | **0.999** | 0.998 |
| **Precision** |  |  |  |  |  |  |
| Positive | 0.752 | **0.903** | 0.856 | 0.827 | 0.589 | 0.856 |
| Equivocal | 0.233 | 0.513 | **0.533** | 0.385 | 0.133 | 0.404 |
| Negative | 1.000 | 1.000 | 1.000 | 1.000 | 1.000 | 1.000 |
| **Recall** |  |  |  |  |  |  |
| Positive | 0.912 | 0.890 | **0.919** | 0.912 | 0.897 | **0.919** |
| Equivocal | 0.465 | 0.465 | 0.372 | 0.465 | **0.767** | 0.442 |
| Negative | 0.997 | **1.000** | **1.000** | 0.999 | 0.991 | 0.999 |

**Table S2:** Individual probability threshold calibration with associated performance based on differential ROC analysis for quality control and diagnosis. The AUC for quality control relates to the performance on the calibration cohort at the given probability threshold for individual tiles containing columnar epithelium on H&E. The AUC for diagnosis relates to the performance on the calibration cohort at the given probability threshold for individual tiles containing positive goblet cells on TFF3. Sensitivity is based on a fixed value of specificity derived from the pathologist performance on the calibration cohort. The tile number threshold is the resulting cutoff from the fixed specificity.

|  | AlexNet | DenseNet | Inception | ResNet | SqueezeNet | VGG |
| --- | --- | --- | --- | --- | --- | --- |
| **Quality control** |  |  |  |  |  |  |
| Probability threshold | 0.97 | 0.96 | 0.995 | 0.96 | 0.85 | 0.99 |
| AUC | 0.985 | 0.984 | 0.986 | 0.986 | 0.980 | **0.988** |
| **Diagnosis** |  |  |  |  |  |  |
| Probability threshold | 0.9999 | 0.87 | 0.655 | 0.93 | 0.99999 | 0.93 |
| AUC | 0.80 | 0.82 | 0.83 | 0.83 | 0.80 | 0.83 |
| Sensitivity at fixed specificity (91.57%) | 63.4% | 62.5% | 61.5% | 63.5% | 60.6% | **64.4%** |
| Tile number threshold | 3 | 8 | 10 | 9 | 4 | 6 |

**Table S3:** Performance of all architectures after application on the validation cohort. Quality control models relied on pathologist calls on sample quality. Sensitivities or specificities were not determined due to irrelevance in the fully automated model approach. Diagnosis models relied on thresholds determined on the calibration cohort.

|  | AUC (CI 95%)<br>vs. pathologist | AUC (CI 95%)<br>vs. endoscopy | Sensitivity<br>(CI 95%) | Specificity<br>(CI 95%) |
| --- | --- | --- | --- | --- |
| **Quality control** |  |  |  |  |
| AlexNet | 0.98 (0.97-0.99) | n/a | n/a | n/a |
| DenseNet | 0.98 (0.97-0.99) | n/a | n/a | n/a |
| Inception v3 | 0.98 (0.97-0.99) | n/a | n/a | n/a |
| ResNet-18 | 0.97 (0.96-0.99) | n/a | n/a | n/a |
| SqueezeNet | 0.97 (0.95-0.98) | n/a | n/a | n/a |
| VGG-16 | 0.99 (0.98-0.99) | n/a | n/a | n/a |
| **Diagnosis** |  |  |  |  |
| Pathologist | n/a | n/a | 81.75%<br>(76.67%-85.92%) | 92.75%<br>(89.37%-95.51%) |
| AlexNet | 0.96 (0.94-0.98) | 0.86 (0.83-0.89) | 72.24%<br>(66.98%-77.37%) | 89.70%<br>(85.80%-92.97%) |
| DenseNet | 0.97 (0.96-0.99) | 0.89 (0.86-0.91) | 70.34%<br>(64.84%-76.24%) | 92.75%<br>(89.84%-95.85%) |
| Inception v3 | 0.97 (0.96-0.99) | 0.88 (0.85-0.91) | 69.96%<br>(64.71%-75.65%) | 93.13%<br>(89.74%-96.03%) |
| ResNet-18 | 0.97 (0.95-0.98) | 0.88 (0.85-0.91) | 72.24%<br>(66.67%-77.18%) | 91.22%<br>(87.72%-94.64%) |
| SqueezeNet | 0.94 (0.92-0.96) | 0.85 (0.82-0.88) | 69.58%<br>(63.59%-74.54%) | 92.37%<br>(88.85%-95.42%) |
| **VGG-16** | **0.97 (0.96-0.99)** | **0.88 (0.85-0.91)** | **72.62%**<br>**(66.72%-77.64%)** | **93.13%**<br>**(89.75%-96.05%)** |

**Table S4:** Characteristics of patients in quality control and diagnosis classes from calibration cohort. For each of the three quality control and diagnosis classes, the number of patients within the class and the paired ground truth is shown.

| **Quality classes** | No confidence | Low confidence | High confidence |
| --- | --- | --- | --- |
| No. of patients | 22 | 27 | 138 |
| Proportion | 11.8% | 14.4% | 73.8% |
| QC positive (path) | 0 | 9 | 137 |
| QC negative (path) | 22 | 18 | 1 |
| **Diagnostic classes** | High conf. negative | Low conf. equivocal | High conf. positive |
| No. of patients | 56 | 59 | 72 |
| Proportion | 30.0% | 31.5% | 38.5% |
| TFF3 positive (path) | 1 | 10 | 71 |
| TFF3 negative (path) | 55 | 49 | 1 |
| Barrett's esophagus | 12 | 26 | 66 |
| No Barrett's esophagus | 44 | 33 | 6 |

**Table S5:** Characteristics of patients in quality control and diagnosis classes from validation cohort. For each of the three quality control and diagnosis classes, the number of patients within the class and the paired ground truth is shown.

| **Quality classes** | No confidence | Low confidence | High confidence |
| --- | --- | --- | --- |
| No. of patients | 55 | 116 | 354 |
| Proportion | 10.5% | 22.1% | 67.4 |
| QC positive (path) | 0 | 35 | 350 |
| QC negative (path) | 55 | 81 | 4 |
| **Diagnostic classes** | High conf. negative | Low conf. equivocal | High conf. positive |
| No. of patients | 145 | 177 | 203 |
| Proportion | 27.6% | 33.7% | 38.7% |
| TFF3 positive (path) | 4 | 33 | 197 |
| TFF3 negative (path) | 141 | 144 | 6 |
| Barrett's esophagus | 18 | 61 | 184 |
| No Barrett's esophagus | 127 | 116 | 19 |

**Table S6:**
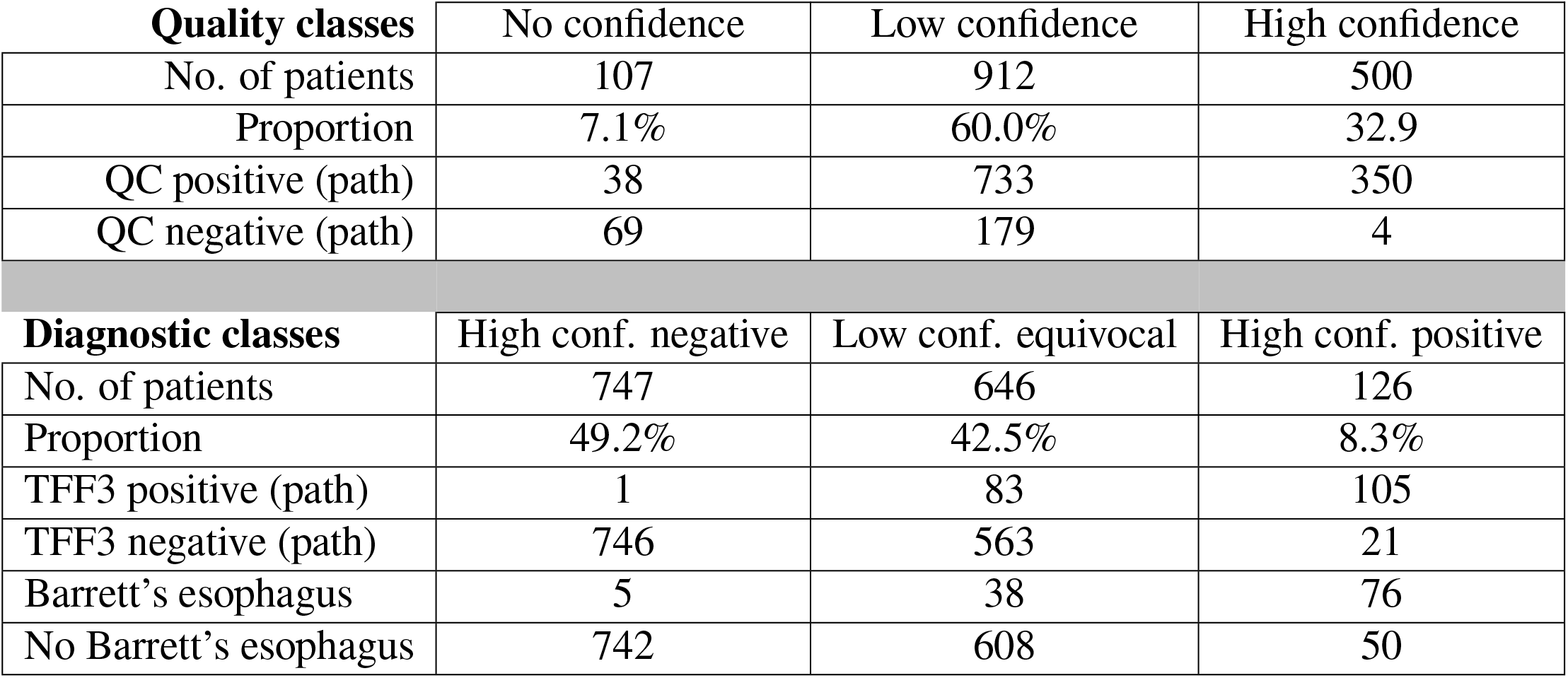
Characteristics of patients in quality control and diagnosis classes from external validation cohort. For each of the three quality control and diagnosis classes, the number of patients within the class and the paired ground truth is shown.

| **Quality classes** | No confidence | Low confidence | High confidence |
| --- | --- | --- | --- |
| No. of patients | 107 | 912 | 500 |
| Proportion | 7.1% | 60.0% | 32.9 |
| QC positive (path) | 38 | 733 | 350 |
| QC negative (path) | 69 | 179 | 4 |
| **Diagnostic classes** | High conf. negative | Low conf. equivocal | High conf. positive |
| No. of patients | 747 | 646 | 126 |
| Proportion | 49.2% | 42.5% | 8.3% |
| TFF3 positive (path) | 1 | 83 | 105 |
| TFF3 negative (path) | 746 | 563 | 21 |
| Barrett's esophagus | 5 | 38 | 76 |
| No Barrett's esophagus | 742 | 608 | 50 |

